# A Southern African Map of Blood Regulatory Variation Enables GWAS Interpretation

**DOI:** 10.1101/2024.09.27.24314510

**Authors:** SE Castel, F Tluway, AK Emde, N Smyth, M Karim, D Sengupta, OA Gray, M Hendershott, S LeBaron von Baeyer, E Burke, S Kaewert, KD Nguyen, SSR Choma, RG Mashaba, LK Micklesfield, C Kabudula, K Kahn, FX Gomez-Olive, S Tollman, A Choudhury, PT Mpangase, S Hazelhurst, KA Wasik, L Yerges-Armstrong, M Ramsay

## Abstract

Functional genomics resources are critical for interpreting human genetic studies, however they are predominantly from European-ancestry individuals. Here we present the Southern African Blood Regulatory (SABR) resource, a map of blood regulatory variation that includes three South Eastern Bantu-speaking groups. Using paired whole genome and blood transcriptome data from over 600 individuals, we map the genetic architecture of 40 blood cell traits derived from deconvolution analysis, as well as expression, splice, and cell type interaction quantitative trait loci. We comprehensively compare SABR to the Genotype Expression (GTEx) Project and characterize the thousands of African-enriched and African-specific regulatory variants mapped. Finally, we demonstrate the increased utility of SABR for interpreting African association studies by identifying putatively causal genes and molecular mechanisms through colocalization analysis of 83 blood-relevant traits from the PAN-UK Biobank. Importantly, we make full SABR summary statistics publicly available to support the African genomics community.

## Introduction

Genome-wide association studies (GWAS) have transformed our understanding of human diseases by revealing underlying biological mechanisms involved in their progression ^1^. This has directly contributed to the development of new diagnostics, precision medicine approaches, and treatments ^2^. However, the vast majority of associations are found in regulatory regions of the genome, making it difficult to determine the causal genes and specific molecular mechanisms, and thus gain novel biological insights ^3^. Functional genomics studies that integrate the genome and multi-omics such as transcriptomics, proteomics, and metabolomics to map molecular quantitative trait loci (QTL) have proven instrumental in bridging the gap between GWAS and mechanisms ^4^. For example, expression QTLs have been proposed to explain a proportion of GWAS heritability, however the exact magnitude has been debated ^5–7^.

To date, the vast majority of GWAS and functional genomics studies have been carried out in European-ancestry individuals, which has contributed to global disparities in our understanding of, and ability to treat disease. As of 2020, almost 90% of all GWAS participants were of European ancestry and over 70% of participants were recruited from just three countries ^8^. While GWAS are becoming more diverse in terms of participant ancestry, a lack of population-scale, ancestrally-aligned functional genomic studies is hindering their translation into actionable discoveries ^9^.

This is particularly true for the African continent, which accounts for over 18% of the world’s population and has the highest amount of human genetic diversity, but has been consistently underrepresented in genomic research ^10–12^. Progress has been made mapping genetic diversity on the continent, however there is still much work to be done ^13–18^. The phenotypic impacts of this genetic diversity are being uncovered through GWAS that include continental African participants, but are still limited ^19–23^. Functional genomic studies that map regulatory variation in continental African cohorts exist but are relatively limited in the cell types captured or have modest numbers of individuals compared to European-ancestry studies ^24–28^.

To help address this African functional genomics gap, we created the Southern African Blood Regulatory (SABR) resource, a map of blood regulatory variation in Southern Africans that includes three South Eastern Bantu-speaking (SEB) groups. Using the same gold-standard methods as the GTEx consortium, we mapped the genetic architecture of blood cell traits derived from deconvolution analysis, as well as expression, splice, and cell type interaction QTLs ^29^. SABR identified thousands of African-enriched QTLs including many conditionally independent eQTLs, and mapped more regulatory variation compared to GTEx whole blood despite a smaller sample size. Finally, we carried out colocalization analysis of SABR QTLs with blood relevant GWAS in African-ancestry individuals from the Pan-UK Biobank and identified both known and novel putatively causal genes and molecular mechanisms for trait associations, demonstrating the utility of the resource to the African genomics community.

## Results

### Whole Genome and Blood Transcriptome Sequencing of Three South Eastern Bantu-speaking Groups in South Africa

Participants were selected for inclusion in SABR from the previously recruited AWI-Gen cohort which had existing genotype data generated using the H3Africa SNP array for 10,603 individuals ^13,19,30^. The AWI-Gen cohort includes older adults randomly selected from established population-based cohorts. They include the Agincourt and DIMAMO health and socio-demographic surveillance systems in rural northern South Africa, and the urban Soweto cohort at the Developmental Pathways for Health Research Unit in Gauteng ^31,32^. These three AWI-Gen research centers recruited participants from eight major South African SEB groups, and three of these groups - Pedi or Bapedi (Sepedi speakers, Limpopo Province), Tsonga (Shangaan ethnic group who speak Xitsonga, Mpumalanga Province), and Zulu (isiZulu speakers, Gauteng Province) were included in this study (Figure 1a). Building on respectful and sustained precedents of community involvement by public engagement teams of the host research centers, participants were selected for inclusion in this study based on clustering closely with one of the three SEB groups in principal component analysis and being minimally related. Extensive site-specific community engagement was carried out to introduce and explain the proposed study to community leaders and potential participants. Participants were invited into the study and following informed consent, 750 were enrolled in the study, and had venous whole blood collected for DNA and RNA extraction and sequencing.

**Figure 1.**
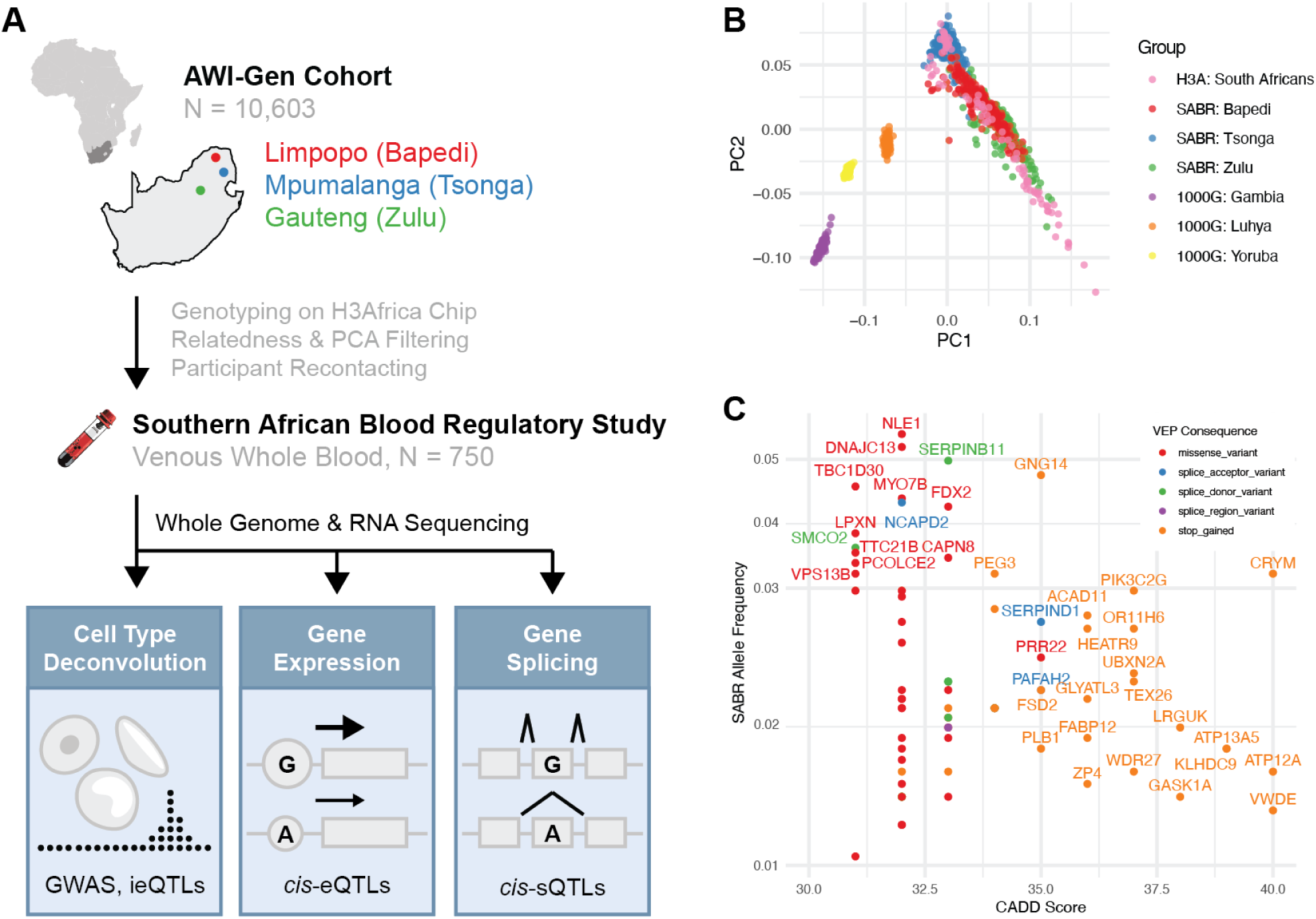
The Southern African Blood Regulatory (SABR) study includes diverse Southern African genetics across three South Eastern Bantu-speaking groups. **A)** SABR blood transcriptome QTL study design. **B)** Principal component analysis of SABR participants and African-ancestry reference groups (1000G = 1000 Genomes, H3A = H3Africa). **C)** CADD score and allele frequency of predicted high-impact variants enriched in at least one of the three South African groups (CADD > 30, AF > 2% and AF > 5x 1000 Genomes Africans in any population).

DNA samples were whole genome sequenced (WGS) to a median coverage of 5.1x (Figure S1a-b, Table S1). Joint genotyping and imputation was performed from WGS data, including relevant reference populations and previously generated high-pass sequencing data of Southern African individuals, using an approach we have previously described that is optimized for mid-pass WGS ^16–18,33,34^. In principal component analysis, Southern Africans formed a distinct cluster from 1000 Genomes Gambian, Luhya, and Yoruba populations, and showed significant variation across individuals (Figure 1b). Analysis within the SABR cohort alone clustered by SEB group (Figure S1c). Given the genetic diversity of the SABR participants, we sought to identify Southern African-enriched functional alleles and found 76 predicted high-impact coding variants (CADD > 30) that were common in at least one SEB group (MAF > 2%) and enriched versus 1000 Genomes Africans (MAF > 5x), including 27 stop-gain variants (Figure 1c, Table S2).

Stranded RNA-sequencing depleting for globin gene transcripts was performed on RNA extracted from venous whole blood, resulting in a median depth of 30M mapped read pairs, and the resulting data showed acceptable quality metrics (Figure S1d, Table S1). In total, 614 samples passed both RNA-seq and genotyping quality control, and were used for QTL mapping. Hidden transcriptome covariates to be included in QTL mapping were inferred using PEER factors ^35^. As expected, they correlated with known biological and technical variables (Figure S2) with PEER factors 1 and 3 being most strongly correlated with RNA integrity number (rho = 0.48, p = 4.30e-7) and DV200 (rho = 0.57, p = 1.16e-53), respectively (Figure S3a-b). Importantly there was no strong separation by study site when clustering by these two factors (Figure S3c-d). RNA-sequencing reads were used to quantify expression levels and intron exclusion ratios of protein coding and long non-coding RNA (lncRNA) genes.

### Cell Type Deconvolution

Whole blood is a highly dynamic tissue composed of many different circulating cell types, the composition of which can change based on both genetics and the environment. While single-cell RNA-sequencing provides the best characterization of blood cell types, it is still cost-prohibitive at population scale. To estimate the relative abundance of relevant blood cell types, we applied cell type deconvolution analysis to the bulk transcriptome data ^36^. Such an approach has previously been used to successfully map cell type interaction eQTLs from bulk data in tissues including whole blood ^37^. Using this approach, we were able to estimate the levels of 40 blood-relevant cell types that had an enrichment score > 0 in at least 50% of individuals (Figure 2a, Table S3). Assuringly, the cell type enrichment score distributions were broadly similar to those derived from GTEx whole blood samples (Figure S4). As expected, numerous PEER factors were correlated with cell type enrichment estimates (Figure S5). PEER factors 1 through 3 were most positively correlated with erythrocytes (rho = 0.57, p = 4.83e-54), neutrophils (rho = 0.64, p = 0), and monocytes (rho = 0.53, p = 0), respectively. Cell type enrichment estimates were also correlated with age and sex, with platelets showing the expected decrease in men (rho = -0.26, p = 7.78e-11) ^38^. At least one cell type was significantly (p < 0.01) correlated with asthma, diabetes, HIV infection, hypertension, obesity, and smoking status, demonstrating the ability of deconvolution to capture disease relevant changes in cell type proportions (Figure 2b, Figure S6). Notably, HIV infection was correlated with decreased levels of regulatory T cells (cor = -0.24, p = 1.22e-8) and CD4+ T cells (cor = -0.16, p = 1.22e-4) and increased levels of CD8+ T cells (cor = 0.27, p = 9.72e-11), replicating known disease biology ^39–41^.

**Figure 2.**
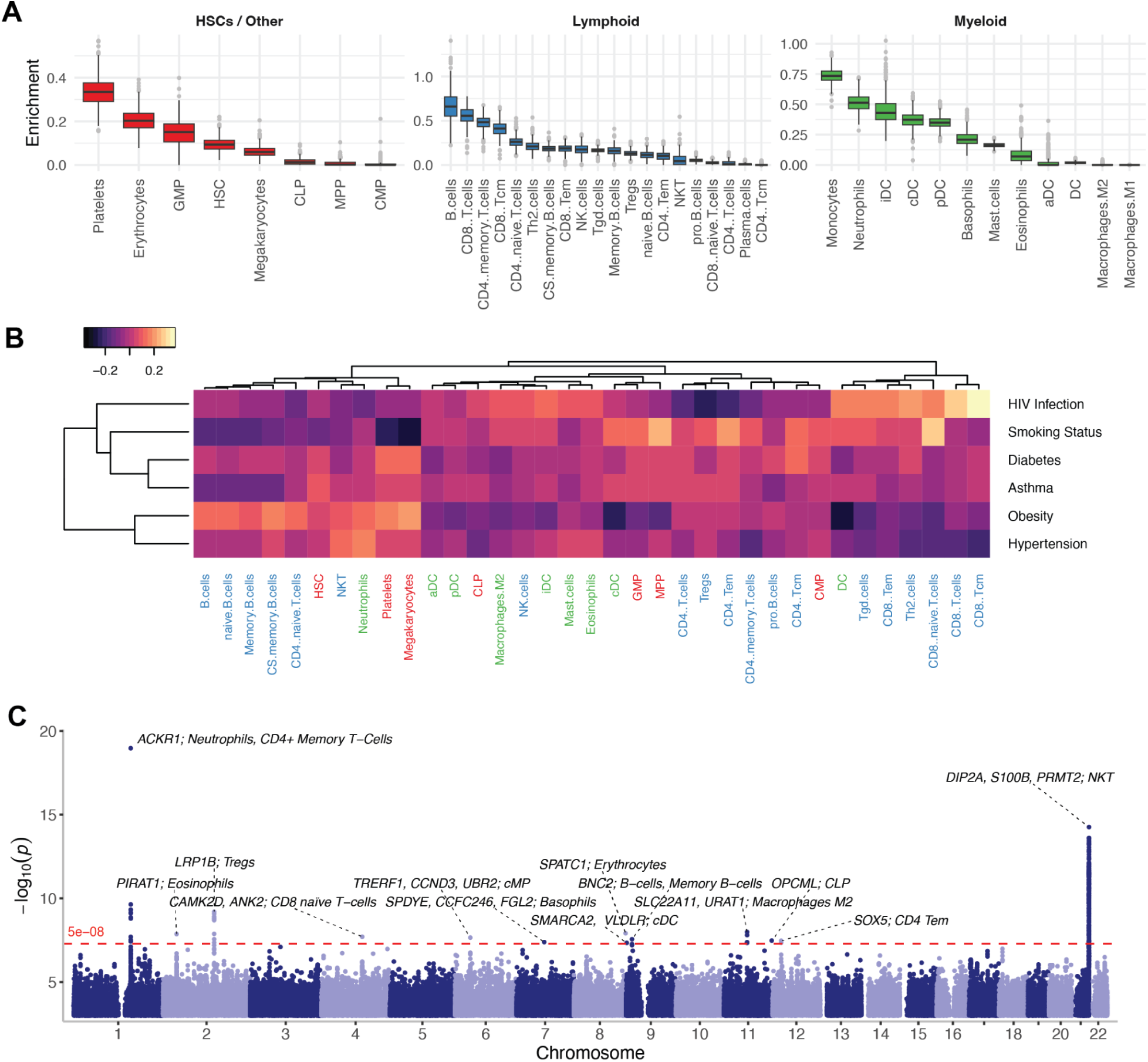
Cell type deconvolution analyses of whole blood transcriptome data. **A)** Distribution of cell type enrichment scores calculated using xCell for 40 blood relevant cell types stratified by lineage - hematopoietic stem cells (HSCs, red), lymphoid cells (blue), myeloid cells (green). **B)** Heatmap of Pearson correlations between binary traits and cell type enrichment scores with cell types labeled by lineage. **C)** Combined Manhattan plot of 19 cell type enrichment GWAS with at least one genome-wide significant hit (p < 5e-8, dotted red line), with nearest genes and cell types listed for each locus.

### GWAS of Deconvoluted Cell Types

The genetic architecture of blood cell counts has proven to be directly relevant to disease biology and therapeutic development, but GWAS have largely been limited to cell types measured in standard complete blood count (CBC) tests ^42,43^. This study presented an opportunity to carry out GWAS of more comprehensive cell types in an underrepresented group. Using the deconvolution estimates we identified 24 loci genome-wide significantly (p < 5e-8) associated with 19 cell types (Figure 2c, Table S4). Importantly, we replicated the well-known association of the *ACKR1* Duffy-null allele with reduced neutrophil counts (rs2814778, p = 1.06e-19, beta = -0.87) ^44^. We also identified novel associations, including between an African-enriched allele and natural killer T cells (rs9979271, p = 5.47e-15, beta = 0.52) and a locus containing an African-enriched missense variant in *SLC22A11* and M2 Macrophages (rs75976740, p = 4.55e-8, beta = -0.48) (Figure S7). In total 6 of 22 unique lead cell type GWAS variants were enriched in Africans versus all other 1000 Genomes continental populations (MAF Africans > 5x all other continental populations), and 3 were enriched in SABR compared to 1000 Genomes Africans (MAF SABR > 5x 1000 Genomes Africans), highlighting the power of diverse cohorts to uncover novel genetic associations for even well-studied traits.

### *cis*-QTL Mapping

We mapped *cis* genetic effects on gene expression (eQTLs) and splicing (sQTLs) for protein coding and long non-coding RNA genes using the same gold-standard approach as the GTEx consortium ^29^. eQTLs were mapped in each of the three SEB groups independently and jointly. The joint approach yielded the highest number of eVariants that were rare in at least one of the three groups (MAF < 1%), the highest estimated number of true positives (Storey’s pi1 = 0.86), and contained the vast majority of genes with a significant eQTL (eGenes) mapped across all four approaches (14,191 of 14,750 = 96%, Figure S8, Table S5), so this approach was used for all analyses moving forward, including for sQTL mapping.

Jointly mapping QTLs in all three SEB groups combined, at least one significant (FDR < 0.05) eQTL was mapped for 14,191 of 16,708 (85%) expressed genes and at least one significant sQTL was mapped for 3,778 of 10,184 (37%) spliced genes (Figure 3a, Table S6-9). The top associated expression (e) and splice (s) variants showed the expected distribution of distance to transcription start site, minor allele frequency, and slope (Figure S9-10). SABR *cis-*QTLs genotyped in GTEx whole blood, which included roughly 12% African American-ancestry individuals, replicated well (eQTLs: Storey’s pi1 = 0.72, slope Spearman rho = 0.81, sQTLs: pi1 = 0.79, rho = 0.87, Figure S11), demonstrating the robustness of *cis-*QTL mapping across cohorts and populations. When accounting for sample size, there was an over 50% increase in the number of genes with significant expression and splice QTLs compared to GTEx whole blood (Figure S12). This does not account for technical differences between the studies, however it does illustrate the advantage of carrying out QTL mapping in a cohort with higher levels of genetic diversity compared to those that are largely of European ancestry.

**Figure 3.**
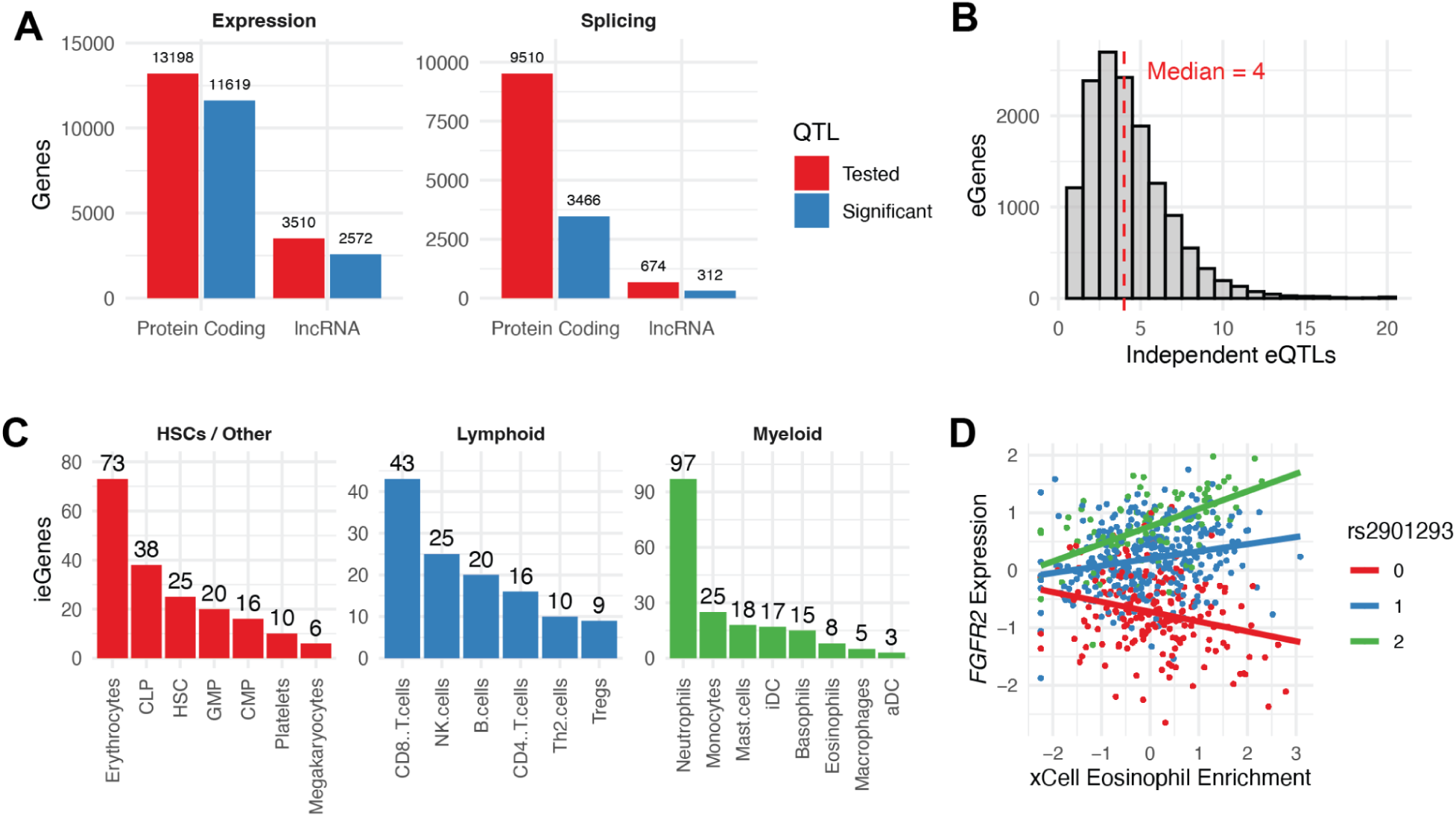
Expression, splice, and cell type interaction *cis-*QTL mapping in whole blood. **A)** Number of genes tested (red) and significant (blue, FDR < 5%) for expression and splice *cis*-QTLs, stratified by gene type. **B)** Number of conditionally independent *cis*-eQTLs mapped per eGene. **C)** Number of significant *cis* interaction eQTLs (ieQTLs) mapped across 21 different cell types stratified by lineage. **D)** Example of a *cis*-ieQTL for *FGFR2* that is dependent on eosinophil enrichment levels (interaction term p-value = 2.22E-13).

### Independent *cis*-eQTL Mapping

African-ancestry individuals exhibit the highest levels of genetic diversity and therefore would be expected to have a high degree of regulatory allelic heterogeneity ^14,16^. This presents an opportunity for QTL studies to map multiple, independent signals per gene. To this end, we carried out independent *cis*-eQTL mapping using a stepwise regression approach and uncovered widespread allelic heterogeneity, with a total of 60,808 conditionally independent *cis*-eQTLs mapped (Table S10-11). There was a median of 4 and mean of 4.3 conditionally independent *cis*-eQTLs per gene (Figure 3b). This compares to a mean of 1.7 *cis*-eQTLs per gene in GTEx whole blood with a roughly equivalent sample size (670 in GTEx vs 614 in SABR), again demonstrating the power of QTL mapping in a genetically diverse cohort.

Conditionally independent eQTLs showed marked differences from primary eQTLs, with decreasing MAF, increasing distance to eGene TSS, and decreasing effect size as a function of conditional index (Figure S13a-d). Furthermore, a number of additional eVariants with predicted high impact were captured by conditional analysis, including 14 stop-gain variants (Figure S13e).

### *cis*-ieQTL Mapping

Cell type interaction eQTLs (ieQTLs) were mapped for 21 cell types that were either medically relevant, or prevalent in whole blood using the same approach as the GTEx Consortium ^37^. This approach includes an interaction term between genotype and deconvoluted cell type score to capture *cis*-eQTLs where the slope is dependent on the estimated enrichment of a given cell type. All 21 cell types had at least one significant ieGene mapped (FDR < 5%), with a total of 499 ieQTLs across 360 genes (Figure 3c, Table S12-13). Mapped ieQTLs included those with cell type specific effects, for example an ieQTL associated with increased expression of *FGFR2* in eosinophils, which is also the blood cell type the gene is most highly expressed in (Figure 3d). Of the 360 ieGenes, 287 were mapped in only one cell type, suggesting that each captured a unique regulatory architecture (Figure S14).

Both this study and Kim-Hellmuth *et al.* mapped neutrophil ieQTLs using the same cell type deconvolution approach in whole blood, which provided an opportunity for replication analysis. However, full summary stats were not available for Kim-Hellmuth *et al.*, so we were unable to test SABR ieQTLs for replication in GTEx. Instead, we tested GTEx ieQTLs for replication in SABR. We note that differences in LD structure could reduce the replication of GTEx ieQTLs in SABR. Nonetheless, we observed replication that was strongly dependent on the significance of the GTEx ieQTL (Figure S15). Replication ranged from strong for the top 150 most significant GTEx ieQTLs (Storey’s pi1 = 0.73, slope Spearman rho = 0.30, p = 2.08e-4) to moderate for the top 800 GTEx ieQTLs (pi1 = 0.25, rho = 0.19, p = 1.10e-7). This shows that the consistent mapping approach between the two studies results in robust replication of the most significant ieQTLs mapped.

### A Map of Southern African Blood Regulatory Variation

As this was the largest blood transcriptome regulatory map generated to date in a continental African population, we sought to characterize the number of African-enriched (more common in continental African populations) or African-specific (only observed in continental African populations) regulatory variants identified. We therefore characterized lead QTL variants as globally common, African-enriched, African-specific, SABR-enriched, and enriched in each separate SABR group (see Supplementary Methods - Variant Annotations for specific definitions). It is important to note that the labels were not mutually exclusive, for example, a variant could be both African-specific and SABR-enriched. Based on these definitions, 35% of all eQTLs mapped were African-enriched and 6.7% were African-specific (Figure 4a). While less substantial as an overall proportion, hundreds of QTLs were SABR-enriched or specific, or enriched in one of the three SABR groups, demonstrating the substantial diversity of regulatory variation across African populations. Examples of African regulatory variation include an African-enriched eQTL for *NIPSNAP3A* (rs34856872, p = 1.35e-61, beta = -1.44) that is a predicted stop-gain variant present at 3.9% frequency in 1000 Genomes African populations and 3.4% in SABR (Figure 4b), and a SABR-specific sQTL for *KIF16B* (rs138620712, p = 3.36e-28, beta = 2.03) that is a predicted splice donor variant absent from 1000 Genomes Africans and present at 2.1% in SABR (Figure 4c). Finally, we found that conditionally independent eQTLs were much more likely to be enriched or specific in Africans, the SABR cohort, and SABR groups as compared to primary QTLs (Figure 4d).

**Figure 4.**
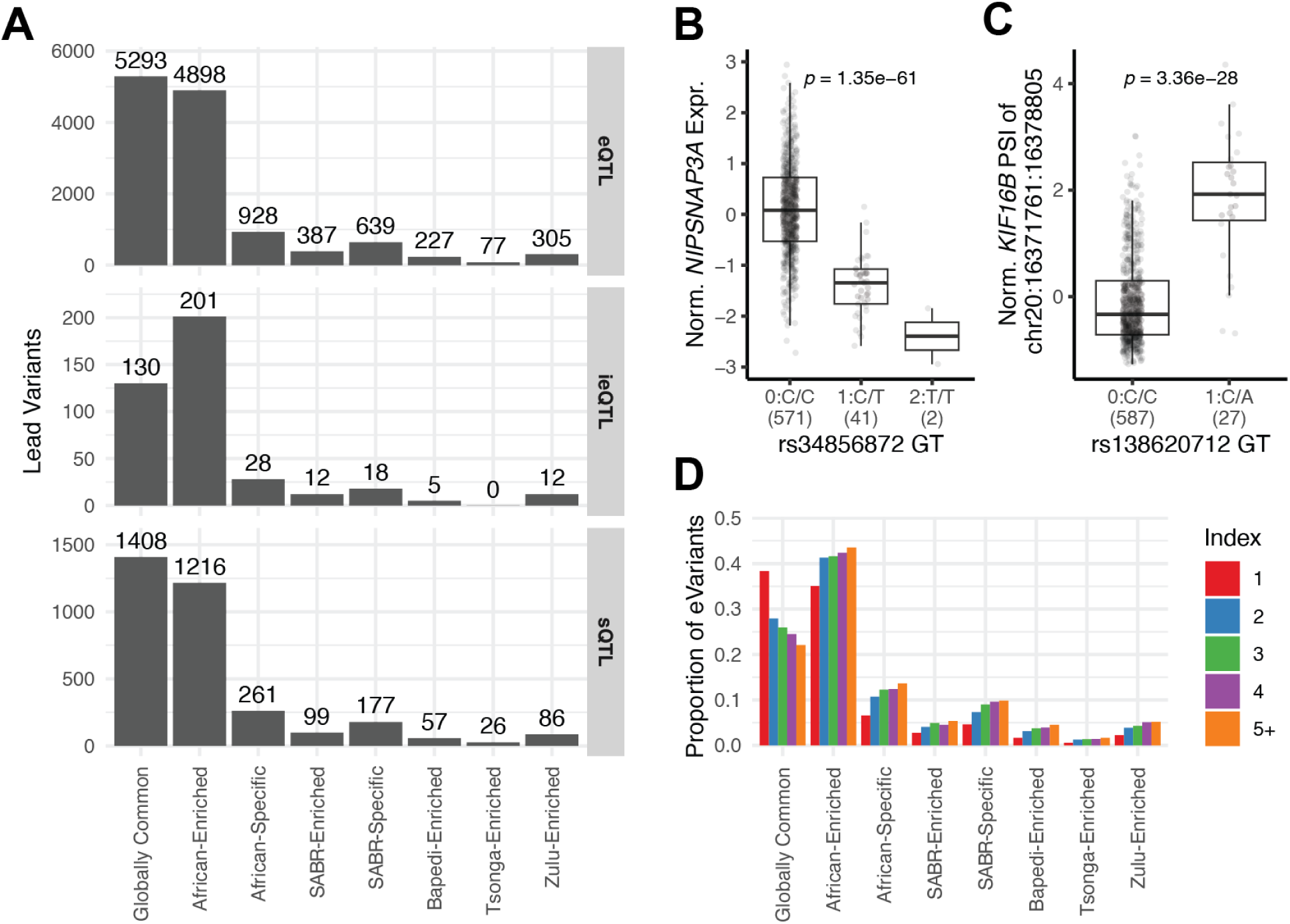
A map of Southern African blood regulatory variation. **A)** Characterization of lead *cis-*QTL variants based on global allele frequencies. **B)** Example of an African-enriched predicted stop-gain eQTL for *NIPSNAP3A* (rs34856872, MAF 1000G Africans = 3.9%). **C)** Example of a SABR-specific predicted splice donor variant sQTL for *KIF16B* (rs138620712, MAF SABR = 2.1%). **D)** Population enrichment and specificity of conditionally independent *cis*-eQTLs stratified by index shown as a proportion of total number of eVariants in each index. Note, for panels **(A)** and **(D)** variants can have more than one label (e.g. African-specific and SABR-enriched). Allele frequency labels are defined in Supplementary Methods - Variant Annotations.

### PAN-UKBB African GWAS Colocalization

To demonstrate the utility of a continental African transcriptome QTL resource for interpreting GWAS, we selected 83 traits where blood is a potentially relevant tissue and performed colocalization analysis with PAN-UK Biobank (PAN-UKBB) GWAS from African-ancestry individuals (Table S14). Given the relatively modest sample size of these GWAS (maximum of 6,636 individuals), we performed colocalization analysis for all loci with a suggestive GWAS signal (p < 5e-6) and a significant QTL (FDR < 5%), at two stringency thresholds (see methods). This resulted in 53 of 83 GWAS traits colocalizing with at least one QTL, and a total of 105 genes colocalizing with at least one GWAS trait, collapsing across QTL types (Table S15).

Analyzing QTL types independently, at the lenient threshold 91 eQTLs, 26 sQTLs, and 5 ieQTLs showed evidence of colocalization (Figure 5a). Collapsing by gene and GWAS trait across QTLs, hematological phenotypes had the highest number of colocalizations, followed by gastroenterology, cardiovascular, and metabolic traits (Figure 5b). Annotating the unique set of lead variants across all loci with evidence of colocalization revealed that 13.8% were entirely absent in European-ancestry individuals (1000 Genomes Europeans MAF = 0) and 30.6% were African-enriched (Figure 5c, Table S16).

**Figure 5.**
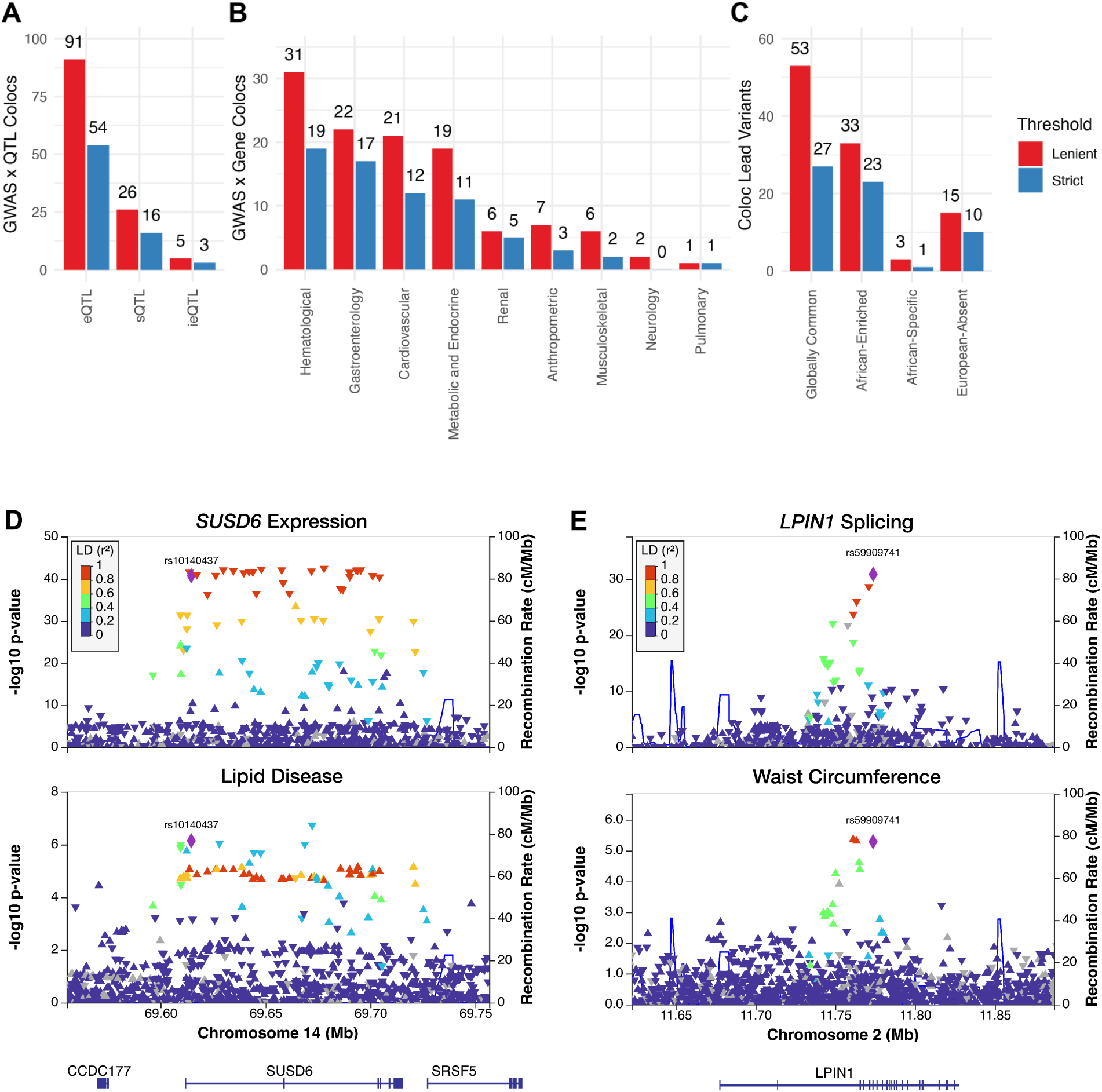
Colocalization of SABR QTLs with PAN-UKBB African GWAS. **A)** Number of colocalizations per QTL type. **B)** Number of colocalizations flattening at the gene-level across QTL types and summarizing based on GWAS trait category. **C)** Characterization of lead colocalization variants based on global allele frequencies. **D-E)** Locus plot of colocalization between an African-enriched *SUSD6* eQTL and lipid disease (**D,** lead variant rs10140437, PP4 = 0.85), and *LPIN1* sQTL and waist circumference (**E**, rs59909741, PP4 = 1.00). In each panel: SABR QTL is shown on top, and GWAS on bottom, lead variant from colocalization analysis is indicated with rsID and set to reference with LD calculated using 1000 Genomes Africans. Allele frequency labels are defined in Supplementary Methods - Variant Annotations. Two thresholds for colocalization were used: lenient (red, PP4 > 0.50 and PP4 / (PP3 + PP4) > 0.80), and strict (blue, PP4 > 0.80).

Many colocalizing African-enriched QTLs were not identified in any GTEx tissue, including an eQTL for *SUSD6* that colocalized with ICD defined lipid disease (rs10140437, PP4 = 0.85) present at 17.5% frequency in 1000 Genomes Africans (Figure 5d) and both an eQTL and sQTL for *C12orf4* that colocalized with serum phosphate levels (rs116827959, PP4 = 0.94) present at 15.4% in 1000 Genomes Africans (Figure S16). Other colocalizing QTLs were not identified in GTEx whole blood, for example a sQTL that colocalized with waist circumference (rs59909741, PP4 = 1.00) present at 23.5% frequency in 1000 Genomes Africans (Figure 5e), or were identified in GTEx whole blood but were orders of magnitude more significant in SABR (Figure S17). This included a stop-gain variant eQTL for *CD36* at 11.5% frequency in 1000 Genomes Africans that colocalized with alkaline phosphatase levels (rs3211938, PP4 = 1.00, Figure S17d). Taken together, these results demonstrate the clear utility of a large, continental African QTL resource for interpreting GWAS in African-ancestry individuals.

## Discussion

As the largest continental African QTL study to date, the Southern African Blood Regulatory (SABR) resource is a small but significant step towards closing the functional genomics gap for global populations. By using a standard approach to data processing and QTL mapping, we have enabled direct comparison of this resource to GTEx, and by making full summary statistics publicly available, it is well placed to become widely used to support genetics research in Africa.

Although SABR does not have the same broad tissue diversity as GTEx, whole blood is nonetheless an ideal tissue for capturing immune function, which is frequently a target of natural selection and shows differences across populations ^45^. Furthermore, whole blood captures a diversity of cell types that can be interrogated through deconvolution analysis, which is not true of transformed cells such as fibroblasts or lymphoblastoid cell lines that are commonly used in QTL studies ^25^. While blood is not always a causal tissue for disease, the majority of *cis*-eQTLs are shared across tissues provided the gene is expressed, and large scale blood eQTL studies have identified increasing numbers of regulatory variants that are active in other tissues ^29,46^.

The SABR resource more extensively explored whole blood as a tissue through analyses of additional deconvoluted cell types compared to previous population-scale studies ^37^. We showed that these additional deconvoluted cell types can capture disease-relevant biology, for example changes in CD4+ and CD8+ T cell levels during HIV infection. However, an important limitation of the approach is that estimates for many cell types have not yet been validated. We also showed that cell type enrichment estimates are suitable traits for GWAS, and can identify novel cell type associations that are not captured by standard complete blood counts. Genetic studies of blood cell traits have uncovered novel biology and contributed to the development of new therapies ^42,43^. This supports carrying out additional deconvolution studies of whole blood transcriptomes in global populations to gain further insight into the genetic architecture of immune traits.

There are both opportunities and challenges when carrying out QTL studies with genetically diverse cohorts. First, it can be challenging to generate high quality genotype data, given that high-pass WGS still remains prohibitively expensive at population scale, and suitable reference panels for genotype imputation are often lacking ^33,47^. Here, we demonstrated that mid-pass WGS offers a cost-effective alternative that is suitable for QTL studies. We also demonstrated that QTL studies with genetically diverse cohorts have the opportunity to map significantly more regulatory variants. For example, SABR mapped 50% more QTLs accounting for sample size versus GTEx whole blood. Further still, despite the smaller sample size, SABR mapped 2.5x the number of conditionally independent eQTLs per gene as compared to GTEx whole blood. The enrichment of population-enriched eQTLs in secondary signals suggests that conditional analyses and large sample sizes should be the standard approach for QTL mapping in diverse populations. Another consideration is that data from admixed individuals can be challenging for standard QTL mapping methods, although approaches have been successfully developed that take into account local ancestry to address this ^48,49^. Finally, QTL resources from populations with smaller haplotype blocks (e.g. African populations) as compared to European-ancestry individuals can enable better fine-mapping of trans-ancestry GWAS signals ^25^.

Even with a limited sample size we uncovered novel cell type associations with potential health-relevance. For example, we identified a locus including a missense variant in *SLC22A11* present at 11% frequency in SABR and absent in other African continental reference populations, that was associated with a transcriptomic signature for M2 macrophages, an immune cell type that is important for the resolution of inflammatory disease. Intriguingly, this locus has been associated with gout, an inflammatory response to monosodium urate crystals in joints, in European-ancestry individuals and M2 macrophages have been proposed to be elevated in chronic gout as a reparative response ^50,51^. *SLC22A11* encodes a urate transporter, thus it is possible that the African-enriched missense variant impacts blood urate levels, gout risk, and thus M2 macrophage levels, although a gout GWAS in a population where this allele is prevalent would be required to determine this. Integrating both cell type GWAS and QTL results also has the potential to uncover novel disease biology. For example, an African-enriched stop-gain variant was identified as a strong *cis*-eQTL for *NIPSNAP3A*. This variant was also nominally associated with increased CD4+ memory T cell levels in cell type deconvolution analysis (p = 4.68e-3), which matches the phenotype observed in mouse knockout studies of the gene ^52^. This human genetic link suggests that NIPSNAP3A may have potential as a target for modifying CD4+ memory T cell levels, which has relevance to autoimmune diseases, cancer, and infectious diseases including HIV/AIDS ^53–55^. It is however important to recognize that the health relevance of trait associations might be different across populations. This is exemplified by Duffy-null associated neutrophil count (DANC), which is phenotypically benign in African-ancestry individuals, demonstrating the importance of carrying out clinical and precision medicine studies with underrepresented groups ^44,56^.

The most common application of QTL resources is to uncover the specific genes and molecular mechanisms underlying genetic associations. By systematically integrating SABR with PAN-UKBB GWAS carried out in African individuals through colocalization analysis, we identified at least one putative causal gene and mechanism for each of over 50 traits. This included identifying novel trait-gene associations. For example, an African-enriched eQTL for *SUSD6* colocalized with lipid disease and the directionality suggested that lower expression is associated with increased risk. This gene has not previously been nominated as a causal gene for lipid disease, however the colocalization agrees with observations from mouse studies, where knockout of *SUSD6* was associated with decreased HDL and increased liver enzyme levels, markers of lipid disease ^52^. Similarly, an African-enriched sQTL for *LPIN1* colocalized with waist circumference, and this gene has not been previously associated with body composition traits. However, it is known to play a role in lipid metabolism and mouse knockout studies have identified multiple lipid and fat distribution phenotypes, making it a plausible causal gene ^52,57^. The strongest evidence for colocalization observed (PP4 = 0.99) was between a *CD36* African-enriched stop-gain eQTL and alkaline phosphatase (ALP) levels. Reduced expression of *CD36* was associated with decreased ALP levels, indicating that loss of function could be protective in the context of liver disease. Indeed, the stop-gain variant is significantly associated with increased HDL levels in GWAS (p = 3.00e-27) and mouse studies have shown the gene is involved in fatty acid metabolism, with disruption in liver protecting from steatosis and improving lipid profiles ^58,59^. Across all colocalizations, 13.8% of lead variants were entirely absent in Europeans and 30.6% were African-enriched, clearly demonstrating the need for African functional genomics resources.

Studies that integrate the genome and omics in diverse populations are becoming increasingly common and include the plasma proteome and metabolome ^60,61^. However, transcriptome studies based on RNA-sequencing still offer the greatest breadth and depth in a cost-effective, single assay due to the untargeted nature of the underlying technology. This allows many molecular traits to be quantified from a single data type, as demonstrated by the cell type deconvolution, expression, and splicing analyses in the SABR resource. Future versions of the resource could be expanded to include additional molecular phenotypes such as alternative transcription start site usage and polyadenylation using the same data ^62^. While blood is most frequently collected for transcriptome studies, non or minimally invasive biospecimens such as hair, urine, or skin biopsy could be used in future studies to explore additional biology ^63^. Finally, multi-omic studies have revealed that data types are often complementary rather than redundant with one another, each capturing a unique aspect of tissue biology ^46^.

Given the vast genetic diversity of the African continent, SABR (Southern Africa) and others like the African Functional Genomics Resource (Western and Eastern Africa) are just a small part of the many functional genomic studies that are required ^25^. This is also true of African genome-wide association studies, which are making progress but still lag behind those carried out with European-ancestry populations. Developing these resources with the input of participating communities and in a way that benefits them is essential and will directly contribute to our understanding of diseases with specific relevance to African people ^64–66^. Together, these will work towards a vision of precision medicine with the potential to improve the health of the billions of people living on the continent.

## Acknowledgements

The study acknowledges the generosity of the participants who shared information, provided blood samples and gave us permission to use and share their data widely with the research community. The community engagement and benefit sharing activities were enabled through the extraordinary efforts of the following people at each of the Research Centres: Jackson Mabasa (DPHRU), Simon Khoza (SAMRC/Wits Agincourt Unit), and Mmalehlokoana Maria Manabile (DIMAMO). The AWI-Gen Collaborative Centre that contributed foundational data for this study was funded by the National Institutes of Health (NIH) (award number U54HG006938) and the South African Department of Science and Innovation (award number DST/CON 0056/2014). The MRC/Wits Rural Public Health and Health Transitions Research Unit and Agincourt Health and Socio-Demographic Surveillance System, a node of the South African Population Research Infrastructure Network (SAPRIN), is supported by the Department of Science and Innovation, the University of the Witwatersrand, and the Medical Research Council, South Africa, and previously the Wellcome Trust, UK (grants 058893/Z/99/A; 069683/Z/02/Z; 085477/Z/08/Z; 085477/B/08/Z). M.R. is a South Africa Research Chair on Genomics and Bioinformatics of African Populations supported by the National Research Foundation of South Africa (Ref Number 89646). This work represents the views of the authors and does not necessarily reflect the views of the funders. It has enabled securing additional NIH funding (award number U01HL172182) for a project led by A.C. and titled ‘Integrated modeLs for Early Risk-prediction in Africa (ILERA)’.

## Author Contributions

M.R., K.A.W., F.T., S.E.C., E.B., and H.M. designed the study. The Agincourt HDSS managed by the SAMRC/Wits University Rural Public Health and Health Transitions Research Unit is led by S.T., X.G.O., C.K., K.K. and S.S.R.C. and R.G.M. from DIMAMO and M.L.K. from the DPHRU were responsible for managing the community engagement, participant enrollment, blood sampling and processing, N.S., F.T. and E.R. managed biobanking, and F.T., N.S., S.L.V.B., E.B., and P.M. carried out the study. S.E.C., A.K.E., M.K., D.S., O.G., S.K., K.N., A.C., S.H., and L.Y.A. performed data analysis. S.E.C., M.R., and L.Y.A., drafted the manuscript, and all authors contributed to writing and reviewing the manuscript.

## Data Availability

Genome-wide significant cell type deconvolution GWAS associations are available on the NHGRI-EBI GWAS Catalog (accession number XXXXX). Full QTL summary statistics are available in a publicly accessible AWS S3 bucket (s3://xxxxx). All gene-level and annotated summary information for QTLs are included as supplementary tables in this manuscript. All data is made available for non-commercial use only. Accession numbers and supplementary tables will be provided upon manuscript publication.

