## Supplementary Materials for "A Southern African Map of Blood Regulatory Variation Enables GWAS Interpretation"

### Supplementary Tables

**Table S1. SABR Participant Metadata and Sequencing Stats.** Metadata for SABR participants including age, sex, SEB group (group). WGS statistics including mean depth (wgs_mean _dp), proportion of genotype calls imputed (wgs_impute_rate), number of non-imputed genotype calls (wgs_non_imputed). RNA QC metrics including RIN (rna_RIN), percentage of fragments over 200bp (rna_DV200), and number of RNA-seq mapped read pairs (rna_mapped_read_pairs). For each participant, the analyses they were included in is defined (in_wgs = genotype only analyses, in_xcell = cell type deconvolution GWAS, in_qtl = in QTL mapping).

**Table S2. Southern African-Enriched Functional Alleles.** Annotated variants that are predicted to have high functional impact (CADD > 30), common in at least one SEB group (MAF > 2%), and enriched versus 1000 Genomes Africans (MAF > 5x).

**Table S3. xCell Cell Types.** Cell type enrichments that are estimated by xCell, with types included in downstream SABR analyses indicated (included_in_analyses).

**Table S4. xCell GWAS Loci.** All loci that were genome-wide significant in at least one xCell GWAS (p < 5e-8) with top GWAS variant by p-value (lead_variant_id) and corresponding annotations listed.

**Table S5. cis-QTL Mapping Summary.** For each QTL mapping run, summary of number of significant tested genes (tested_genes), QTL genes (FDR < 5%, sig_genes), unique lead QTL variants (unique_lead_variants), tested_genes with minor allele count >= 10 (tested_genes_MAC10), significant QTL genes with MAC >= 10 (sig_genes_MAC10), unique lead QTL variants with MAC >= 10 (unique_lead_vars_MAC10), and minor allele frequency (MAF) in each SEB group, or the entire SABR cohort (ALL).

**Table S6. cis-eQTL Results.** Gene-level permutation results from eQTL mapping using fastQTL.

**Table S7. eVariant Annotations.** Annotations for all unique, lead variants for significant eQTLs (eVariants).

**Table S8. cis-sQTL Results.** Gene-level permutation results from sQTL mapping using fastQTL.

**Table S9. sVariant Annotations.** Annotations for all unique, lead variants for significant sQTLs (sVariants).

**Table S10. Independent cis-eQTLs.** Gene-level results from conditional eQTL analysis, with nominal p-value threshold used for conditional analysis (p_value_theshold), number of conditionally independent eQTLs mapped (independent_signals), and variant IDs (variant_ids). Includes a flag for if an error occurred causing conditional analysis to be stopped before hitting 20 independent eQTLs (error).

**Table S11. Independent eVariant Annotations.** Annotations for all unique, lead variants for conditionally independent eQTLs.

**Table S12. cis-ieQTL Results.** Gene-level results from ieQTL mapping using fastQTL with multiple testing correction using eigenMT with number of independent tests estimated by eigenMT (num_eigenmt_tests) and ieQTL p-value corrected for this number of tests (pval_eigenmt).

**Table S13. ieVariant Annotations.** Annotations for all unique, lead variants for significant ieQTLs (ieVariants).

**Table S14. PAN-UKBB African GWAS.** Summary information for GWAS used in colocalization analysis including trait category (phenotype_category), regression type (model), number of genome-wide significant hits (hits), number of loci with at least one genome-wide significant hit (loci), and full path to summary statistics (path_sumstats).

**Table S15. PAN-UKBB Colocalization Results.** Results with PP4 > 0.50 from colocalization analysis including variant explaining maximum proportion of PP4 (coloc_varant), proportion of H4 explained by coloc_variant (coloc_variant_H4), QTL beta (beta_qtl), GWAS beta (beta_gwas), QTL nominal p-value (pvalue_qtl), and GWAS p-value (pvalue_gwas).

**Table S16. PAN-UKBB Colocalization Variant Annotations.** Annotations for all unique coloc_variants from colocalizations with PP4 > 0.50.

### Supplementary Figures

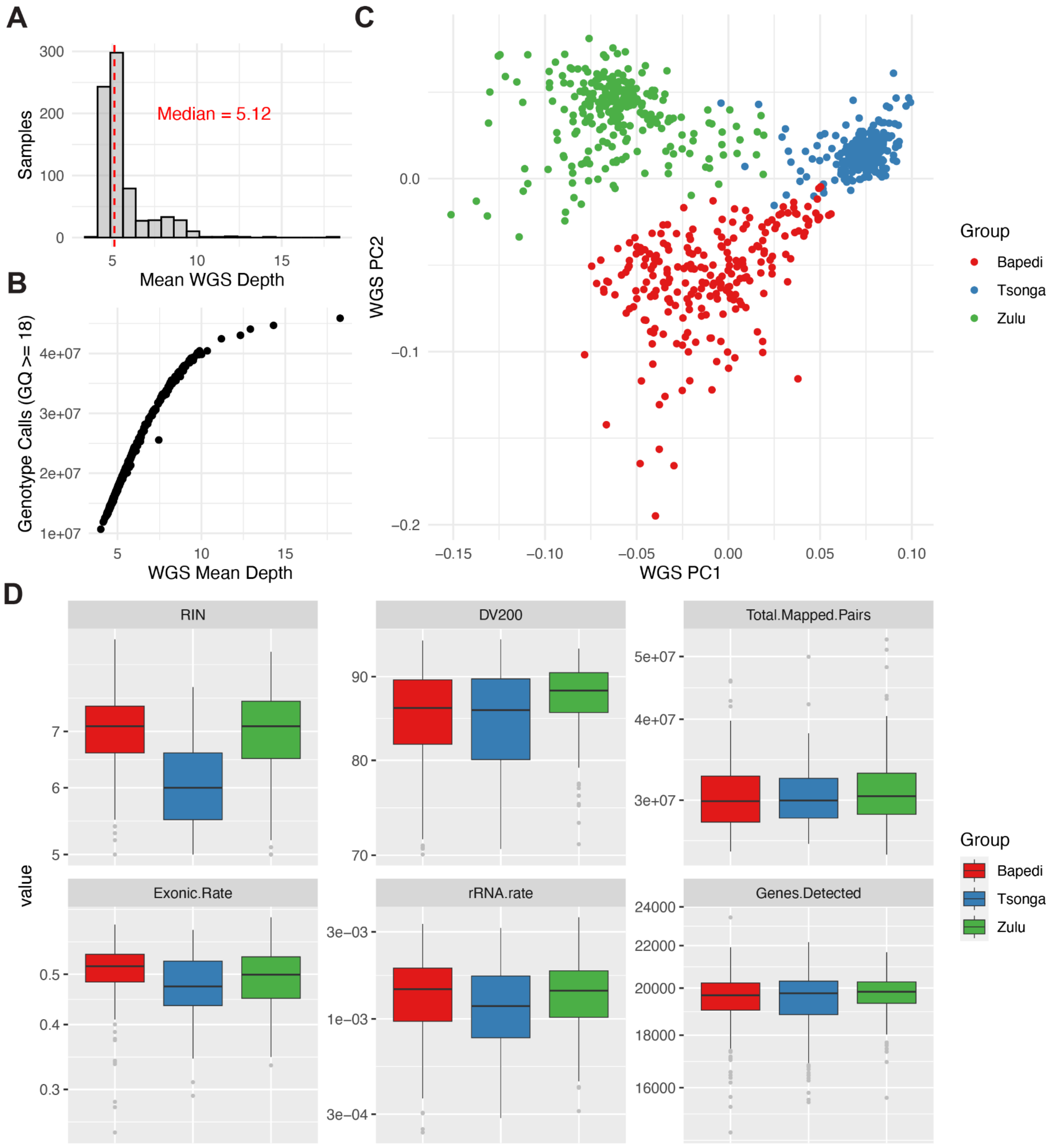

**Figure S1. Whole genome and blood transcriptome sequencing of three South Eastern Bantu-speaking groups. A)** Histogram of mean depth achieved in whole genome sequencing. **B)** Number of genotype calls passing quality filtering (GQ >= 18) as a function of mean depth for each sample. **C)** Principal component analysis of SABR WGS samples colored by group. **D)** Quality metrics for RNA samples (RIN, DV200), and RNA-sequencing (total mapped pairs, exonic rate, rRNA rate, genes detected) stratified by group.

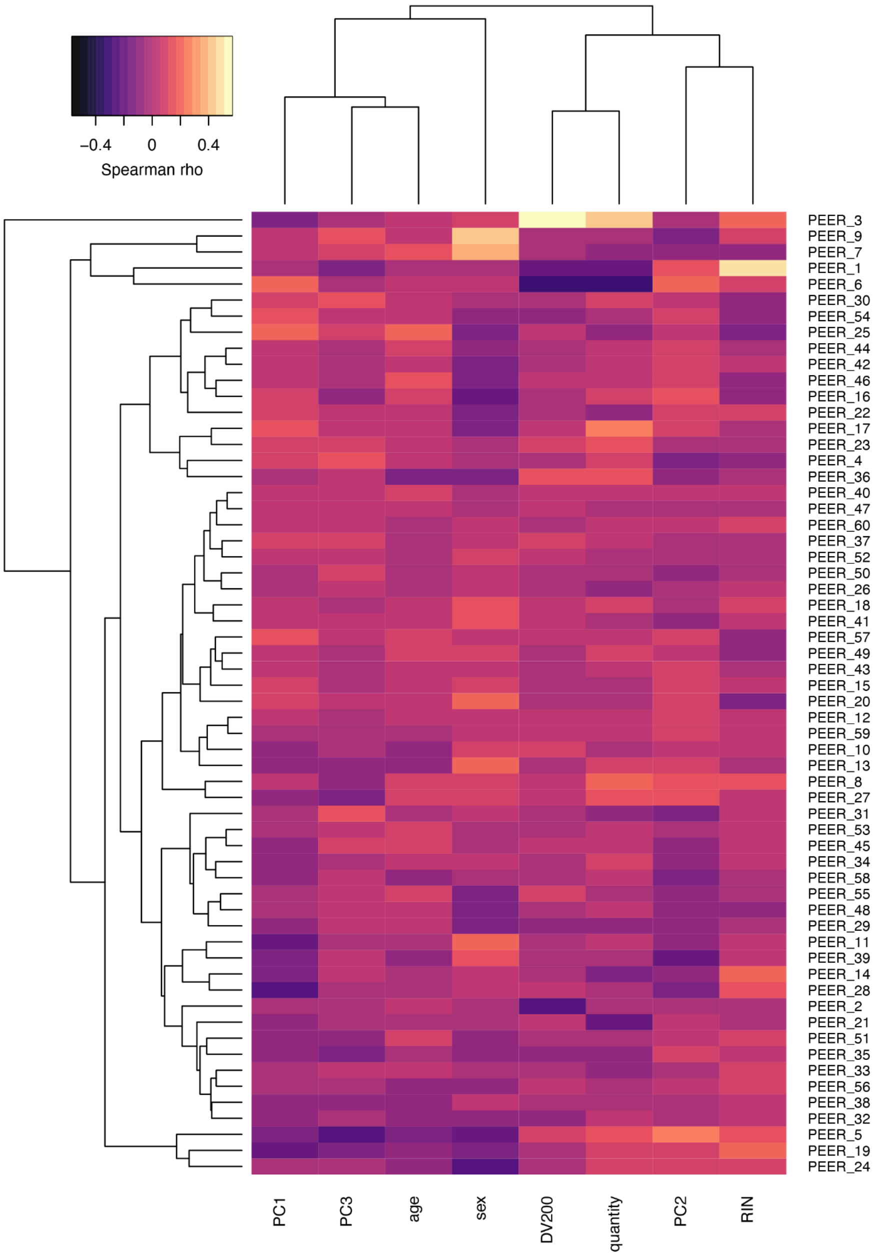

**Figure S2. Heatmap of Spearman correlation between PEER factors and covariates.** PEER factors and covariates have been clustered using euclidean distance. Quantity = quantity of RNA used for library preparation, RIN = RNA integrity number, DV200 = percentage of fragments > 200 nucleotides, PC1-3 = principal components from genotype data.

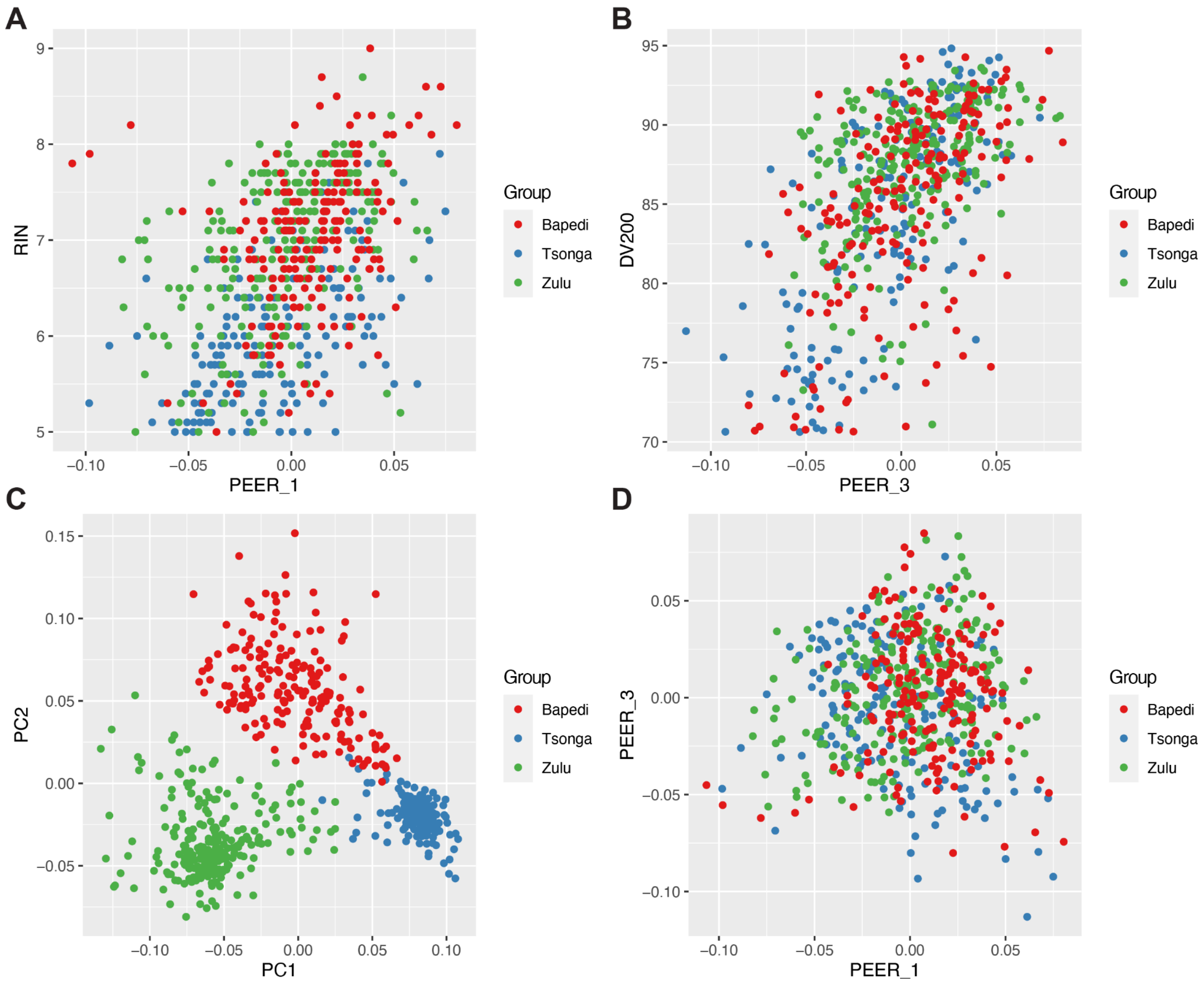

**Figure S3. Correlation of select PEER factors with group, RIN, and DV200. A)** PEER 1 versus RIN, which it is most strongly correlated with. **B)** PEER 3 versus DV200, which it is most strongly correlated with. **C)** PC1 vs PC2 from genotype data. **D)** PEER 1 vs PEER 3 from RNA-seq data. All plots are labeled by SEB group.

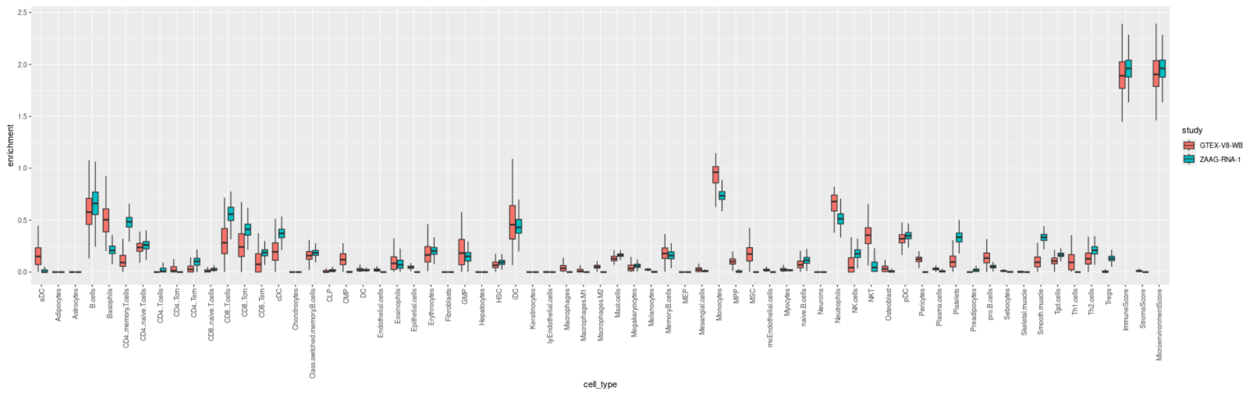

**Figure S4. xCell enrichment score distribution.** Shown for SABR (labeled ZAAG-RNA-1, teal) versus GTEx v8 (labeled GTEX-V8-WB, salmon) whole blood. Outliers are hidden for ease of viewing.

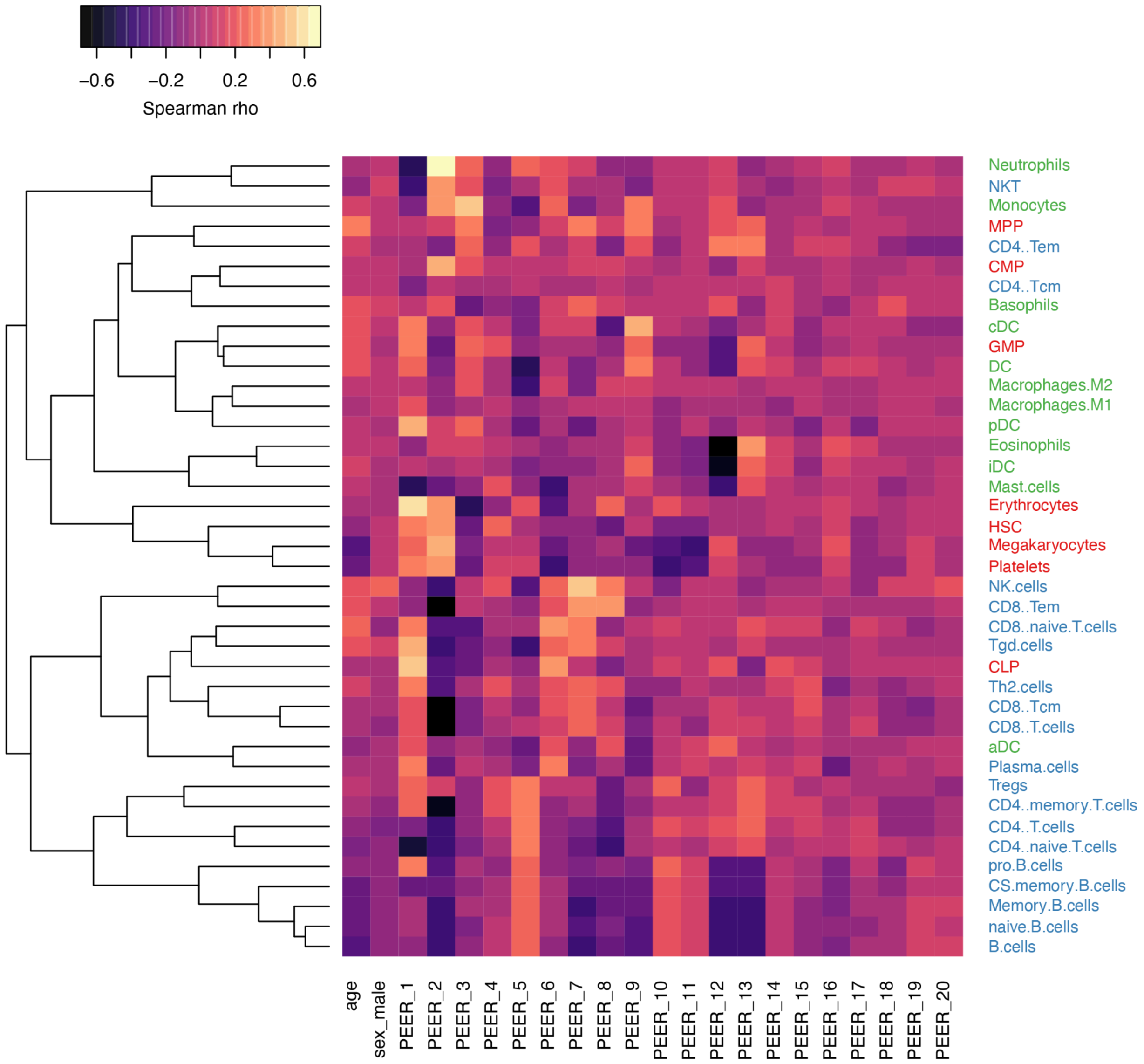

**Figure S5. Heatmap of Spearman correlation between age, sex, PEER factors 1-20 and xCell enrichment scores.** xCell types are colored by group (hematopoietic stem cells: red, lymphoid: blue, myeloid: green). Cell types have been clustered using euclidean distance.

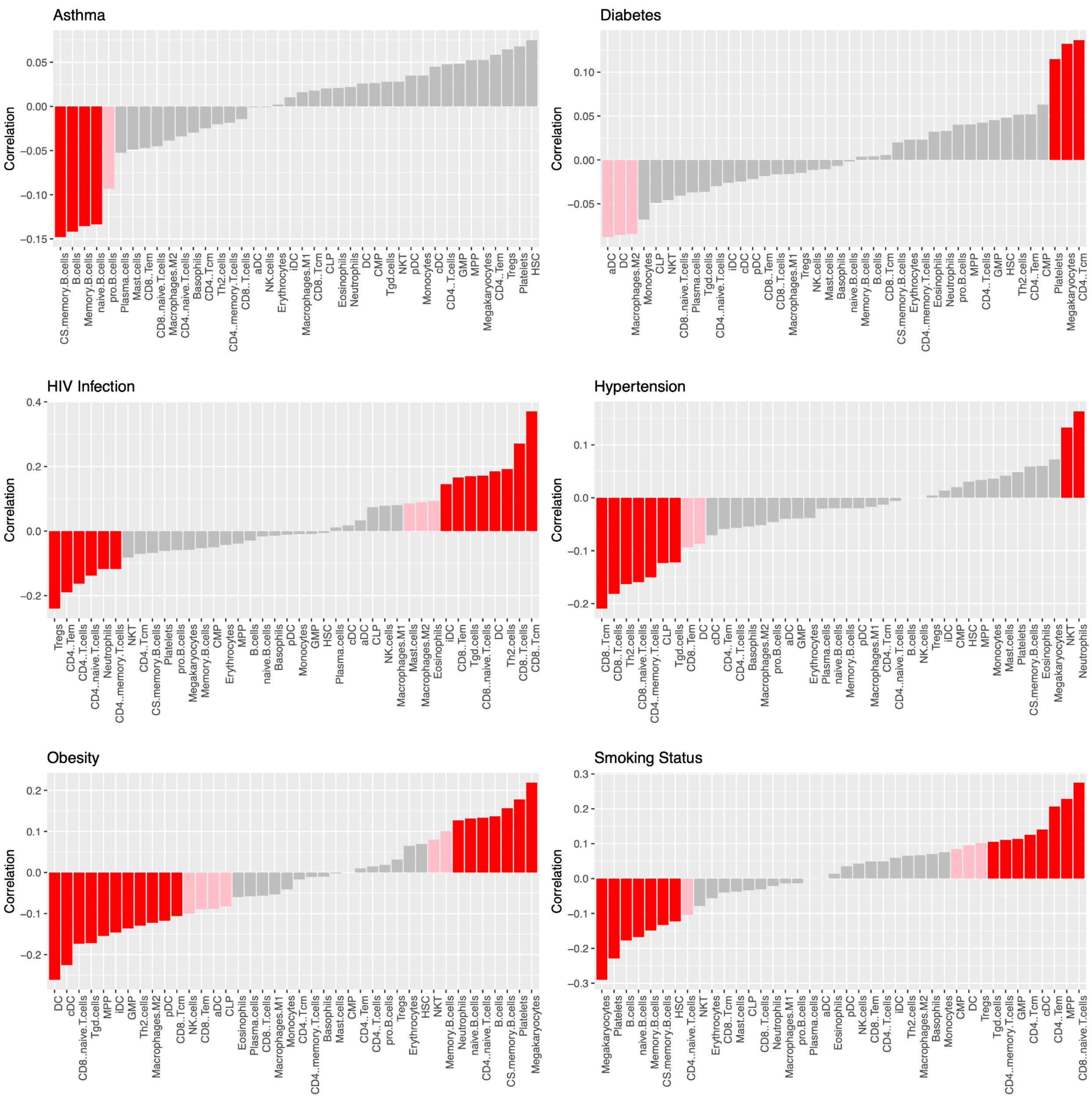

**Figure S6. Pearson correlations between binary traits and xCell enrichment scores.** Correlations bar plots are split by trait (asthma cases = 32, diabetes cases = 31, HIV infection cases = 130, hypertension cases = 296, obesity cases = 214, smokers = 200). Bar plots are colored by statistical significance of the Pearson correlation ($p < 0.01$: red, $0.01 < p < 0.05$: pink, $p > 0.05$: gray).

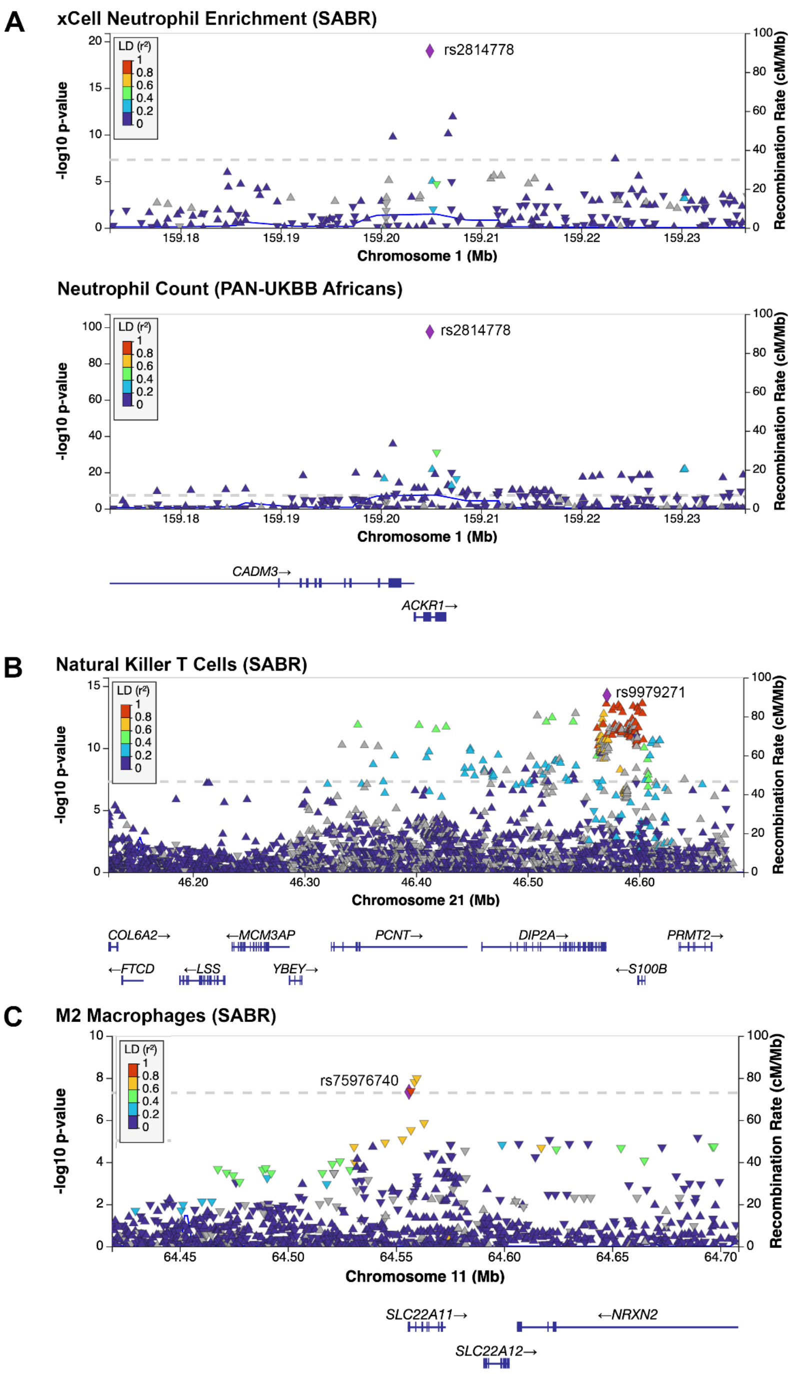

**Figure S7. Locus plots for select cell type deconvolution GWAS. A)** *ACKR1* locus showing associations to neutrophil enrichment score derived from xCell in the SABR cohort (top) and PAN-UKBB neutrophil counts in Africans (bottom), with LD reference set to Duffy-null allele (rs2814778). **B)** Locus associated with xCell natural killer T cell enrichment score in the SABR cohort with LD reference set to lead variant (rs9979271). All locus plots show LD calculated in 1000 Genomes Africans. **C)** Locus associated with xCell M2 Macrophage enrichment score in the SABR cohort with LD reference set to African-enriched *SLC22A11* missense variant (rs75976740). All locus plots show LD calculated in 1000 Genomes Africans.

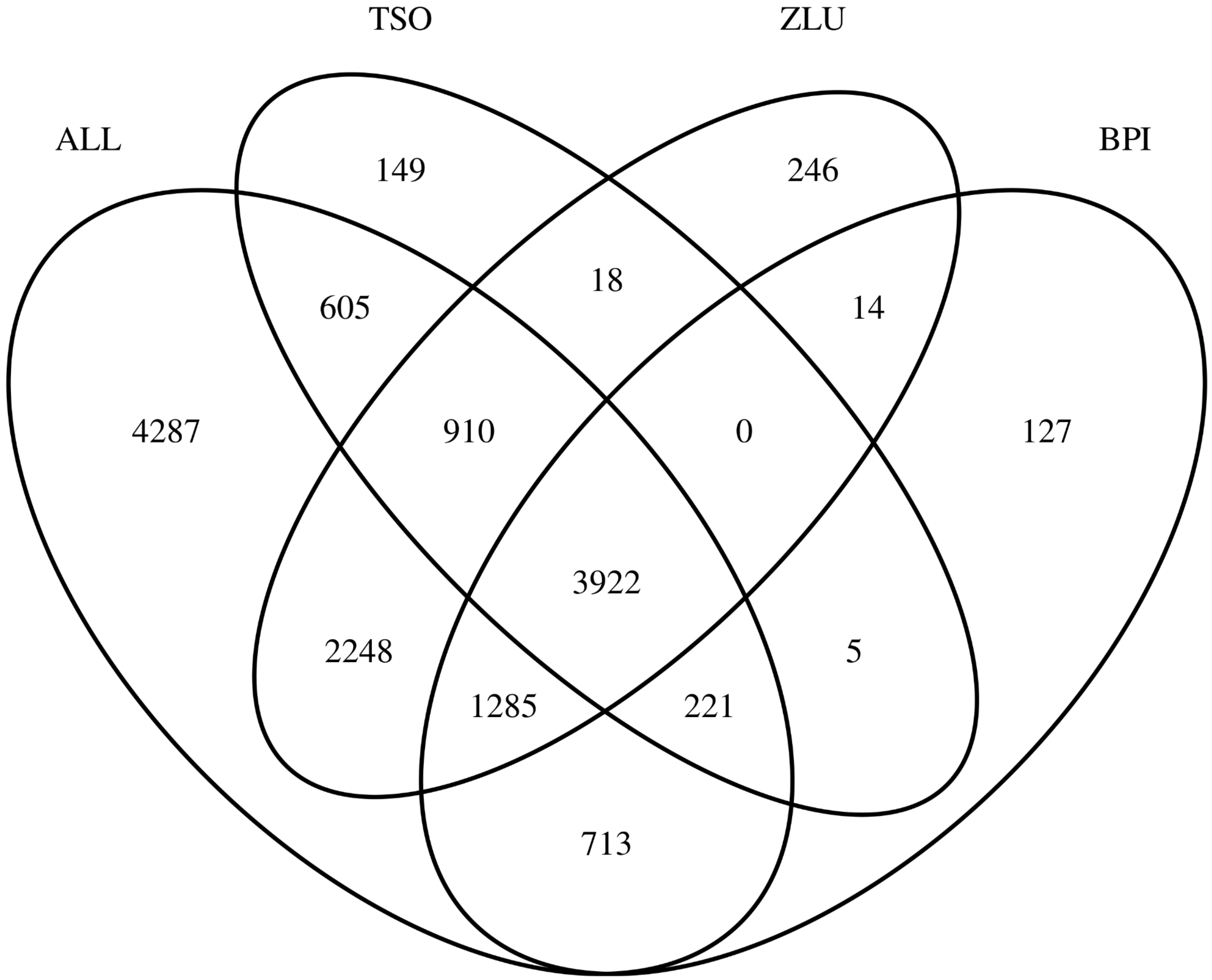

**Figure S8. Venn diagram of significant eGenes mapped across approaches.** Comparison using All SEB groups (ALL, N = 614), Bapedi only (BPI, N = 192), Tsonga only (TSO, N = 181), or Zulu only (ZLU, N = 241). Only eGenes with FDR < 0.05 and eVariant minor allele count >= 10 were included.

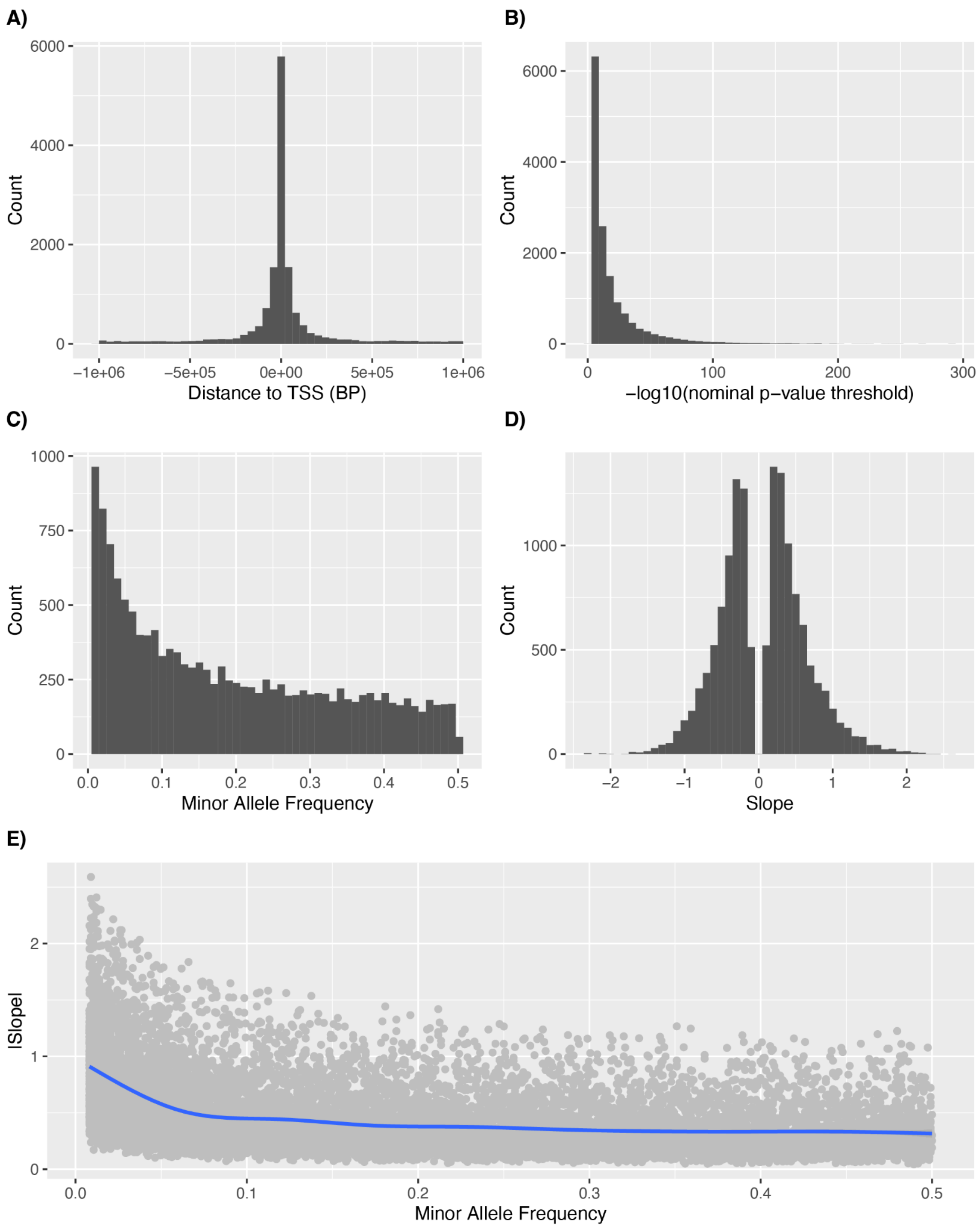

**Figure S9. SABR eQTL summary. A)** Histogram of distance between eVariant and gene transcription start site (TSS). **B)** Histogram of nominal p-value threshold from permutation mapping per eGene. **C)** Histogram of eVariant minor allele frequency in the SABR cohort. **D)** Histogram of eQTL slope. **E)** Scatterplot of eVariant minor allele frequency versus absolute eQTL slope with LOESS regression line (blue). Only significant eQTLs were included (FDR < 0.05).

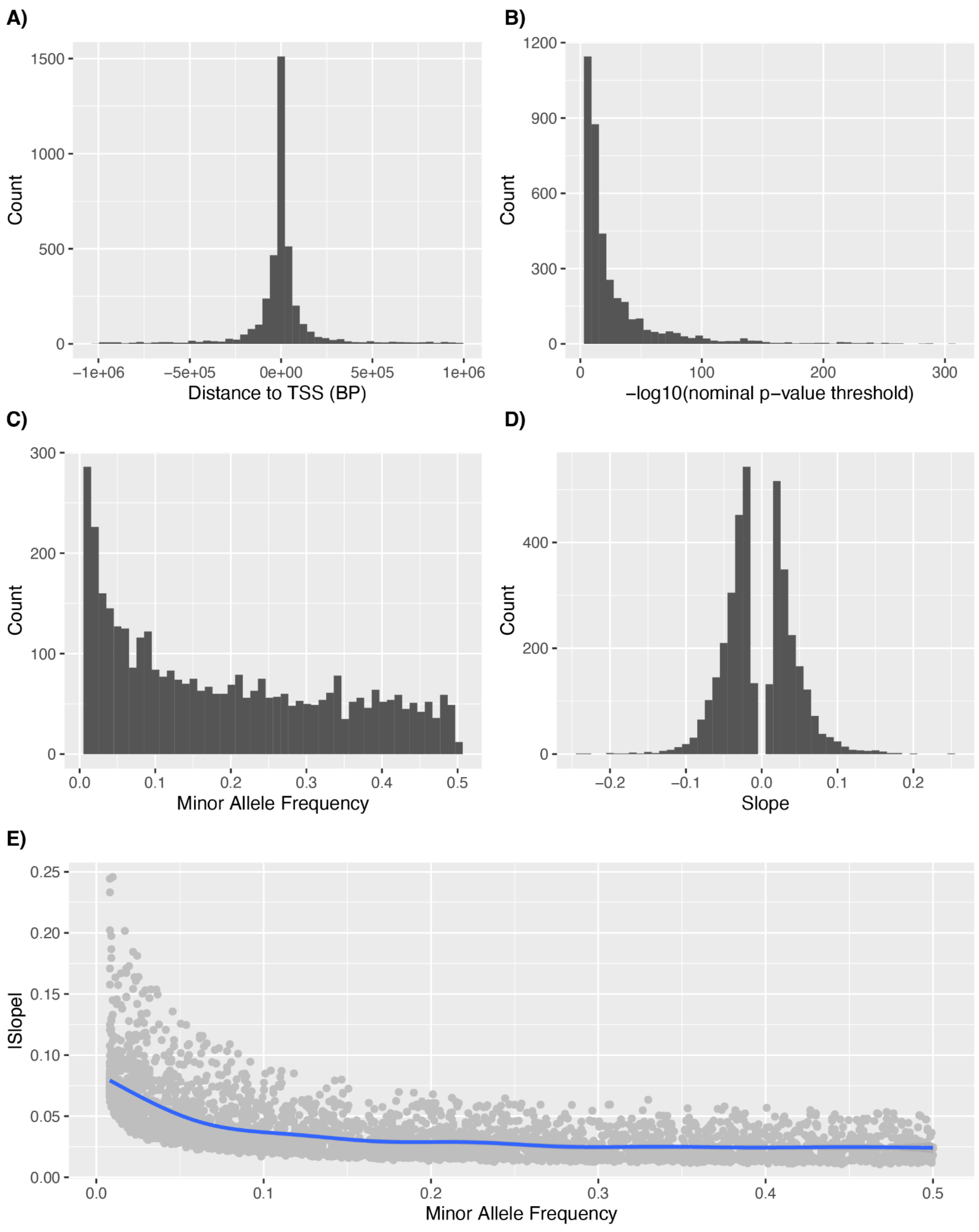

**Figure S10. SABR sQTL summary. A)** Histogram of distance between sVariant and gene transcription start site (TSS). **B)** Histogram of nominal p-value threshold from permutation mapping per sGene. **C)** Histogram of sVariant minor allele frequency in the SABR cohort. **D)** Histogram of sQTL slope. **E)** Scatterplot of sVariant minor allele frequency versus absolute sQTL slope with LOESS regression line (blue). Only significant sQTLs were included (FDR < 0.05).

**A)**

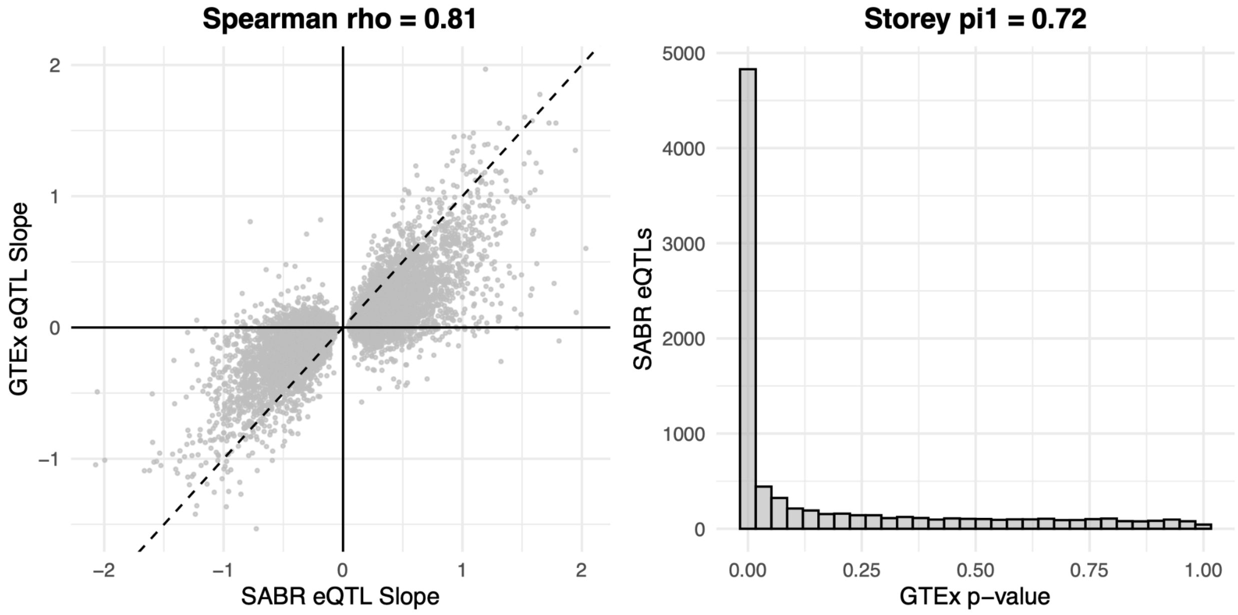

**B)**

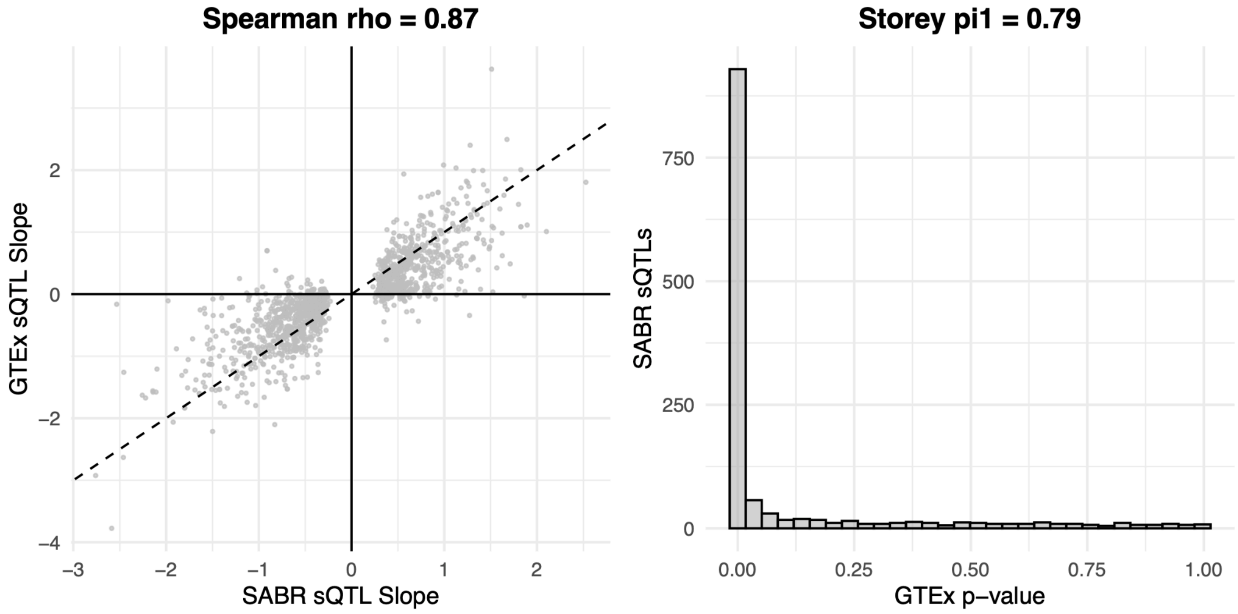

**Figure S11. Replication of SABR *cis*-QTLs in GTEx whole blood. A)** For all SABR eVariants with minor allele count >= 10 in GTEx whole blood (n = 8,610), scatterplot of eQTL slope with Spearman correlation and equality line (left), and histogram of GTEx p-value with Storey's pi1 (right). **B)** For all SABR sVariants with minor allele count >= 10 in GTEx whole blood (n = 1,295), scatterplot of sQTL slope with Spearman correlation and equality line (left), and histogram of GTEx p-value with Storey's pi1 (right). sQTLs were matched based on gene and coordinates of the splicing feature.

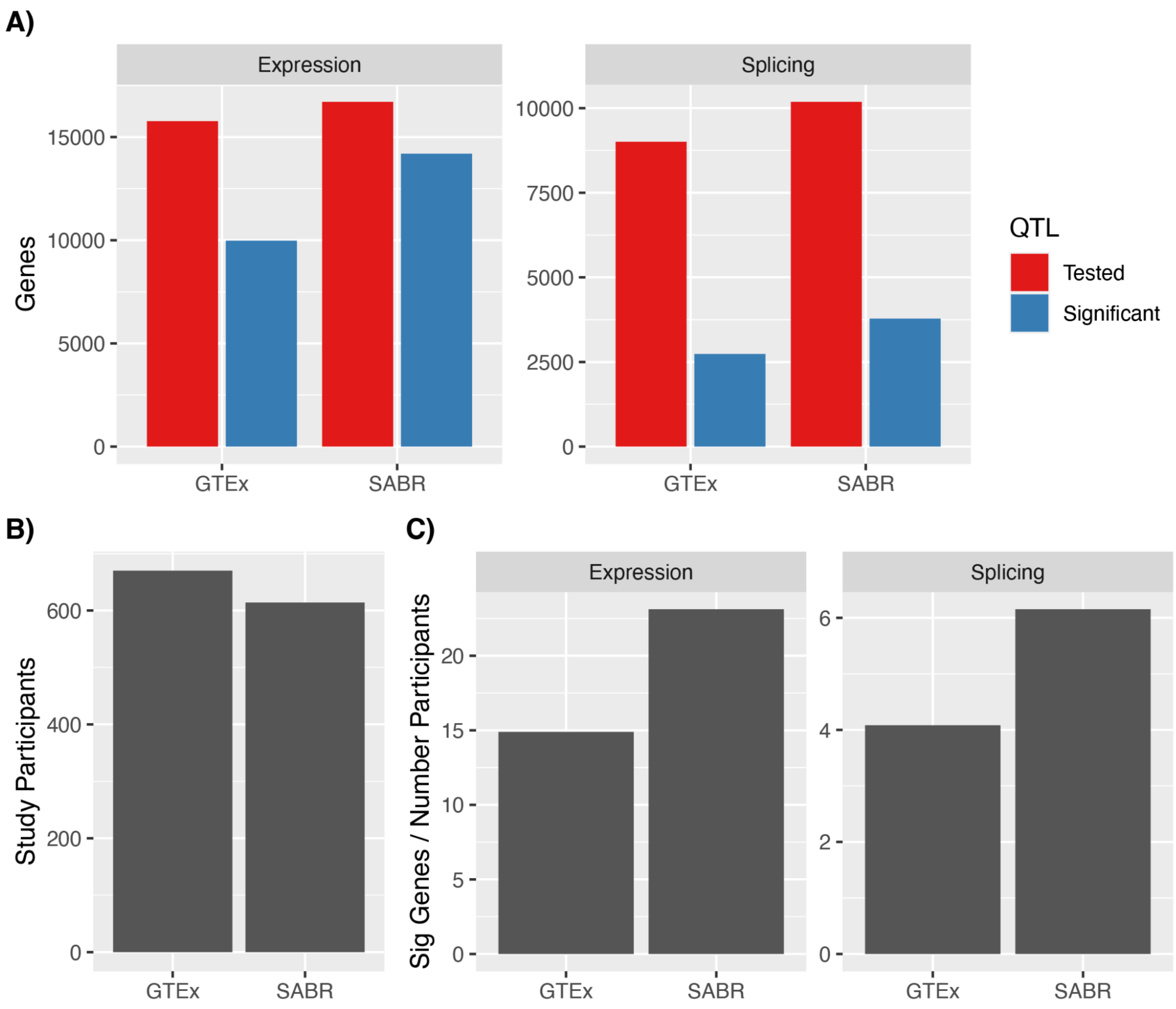

**Figure S12. Comparison of QTL mapping in SABR versus GTEx whole blood. A)** Number of genes tested and those with at least one significant QTL (FDR < 0.05) for expression (left) and splicing (right). **B)** Number of study participants. **C)** Number of genes with at least one significant QTL divided by the number of study participants for expression (left) and splicing (right).

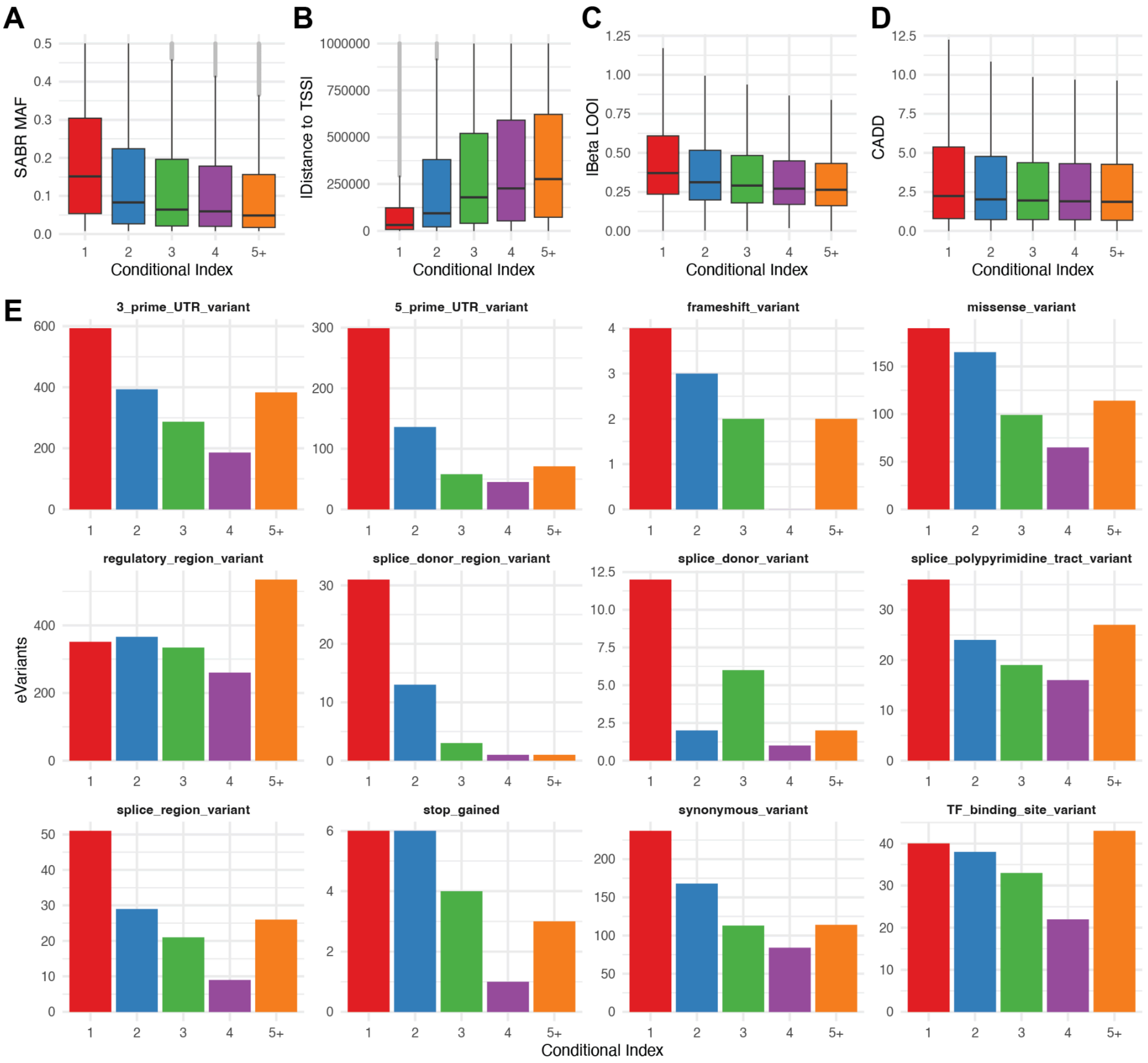

**Figure S13. Functional annotation of conditionally independent eQTL lead variants. A-D)** Boxplot of MAF, absolute distance to eGene TSS, genotype beta from leave-one-out regression, and CADD score for lead eVariants as a function of conditional index. **E)** Count of conditional eVariant predicted effects as a function of conditional index for 12 select effects.

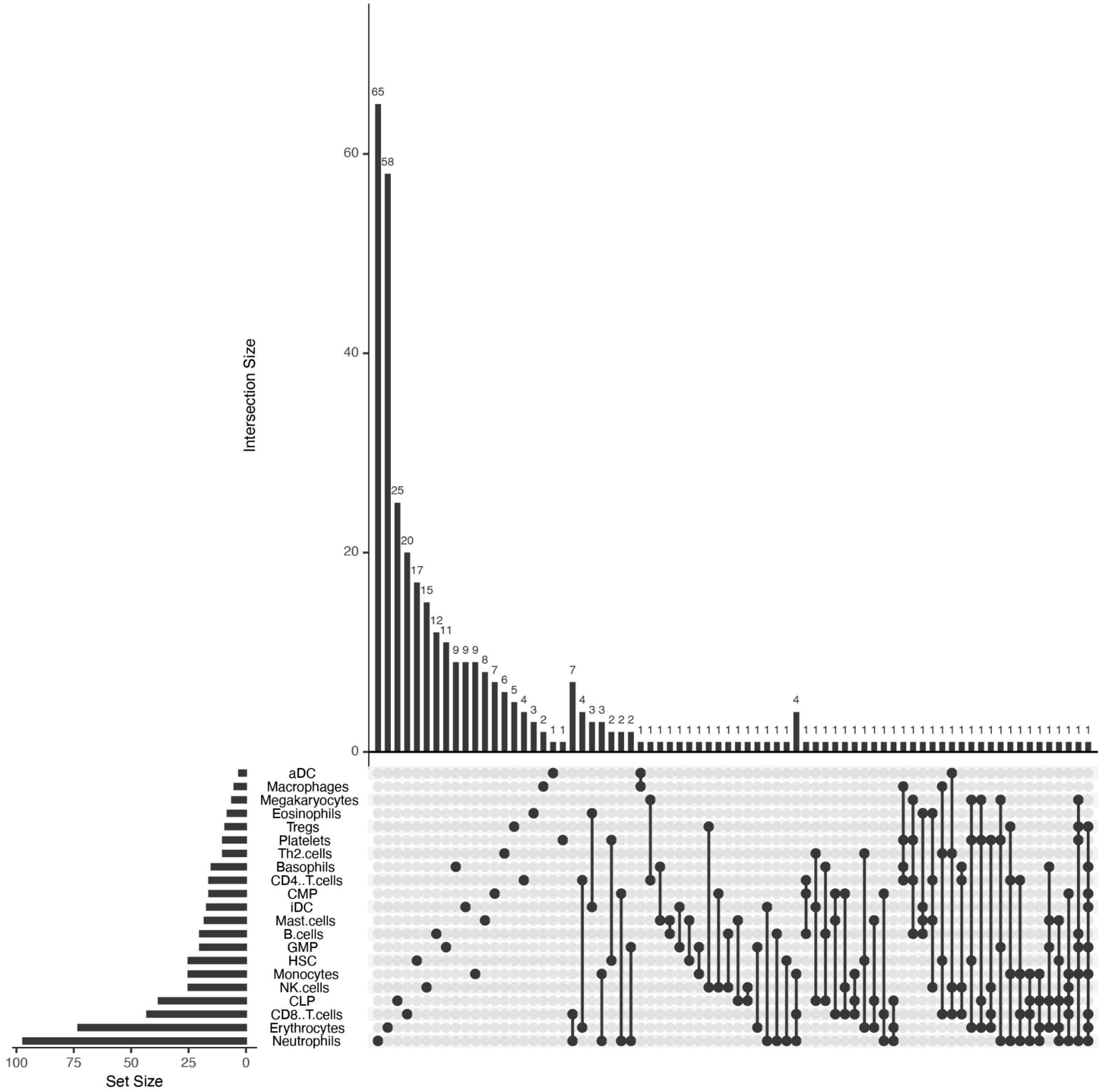

**Figure S14. Intersection of ieQTLs across the 21 xCell types tested.** Vertical bars show the number of ieGenes corresponding to the intersection indicated by the dot plot. Horizontal bars show the number of ieGenes per xCell type.

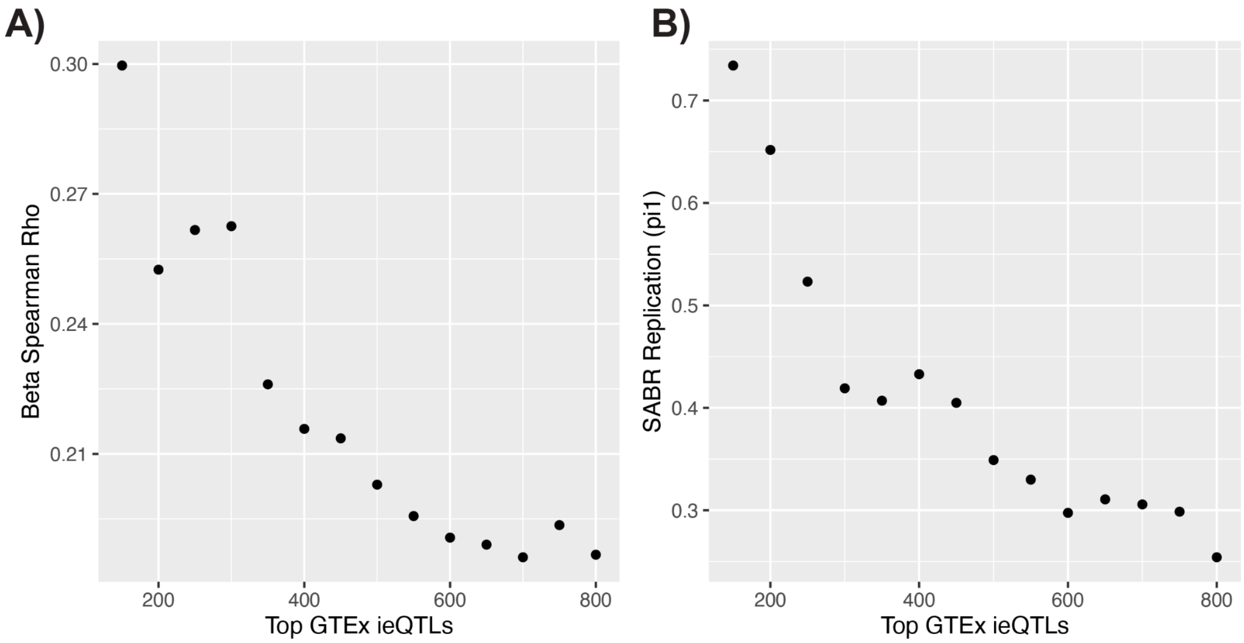

**Figure S15. Replication of GTEx neutrophil ieQTLs in SABR. A)** Spearman correlation of ieQTL interaction term betas as a function of the number of GTEx ieQTLs included, sorted by significance of the ieQTL interaction term in GTEx. **B)** Replication calculated using Storey's pi1 statistic as a function of the number of GTEx ieQTLs included. Only ieQTLs with FDR < 5% in GTEx and MAF > 5% in both GTEx and SABR were included in the analysis.

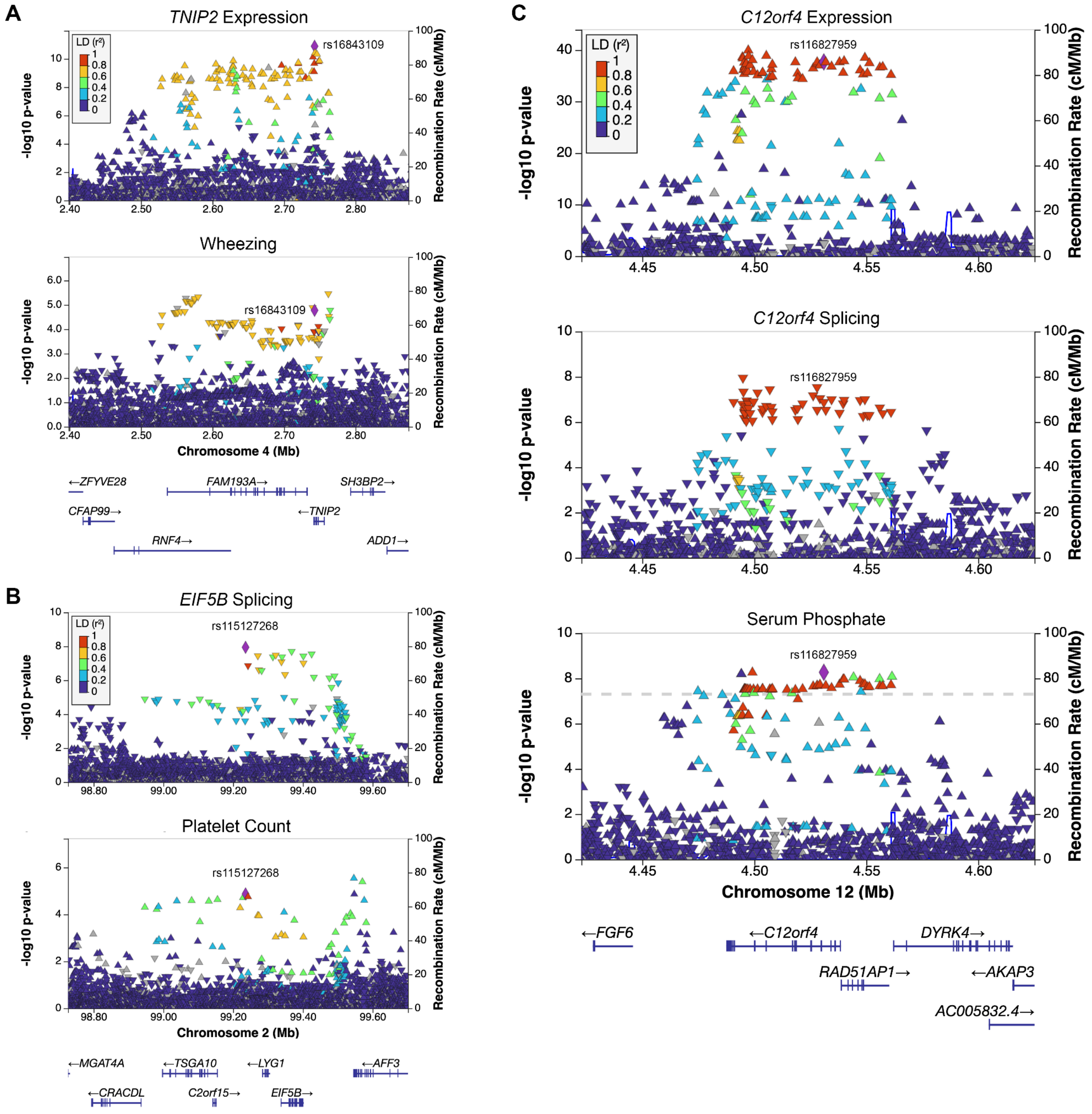

**Figure S16. PAN-UKBB African GWAS colocalizations with African-enriched SABR QTLs not captured in any GTEx tissue**. **A)** An eQTL for *TNIP2* colocalizes with wheezing (rs16843109, PP4 = 0.87). **B)** A sQTL for *EIF5B* colocalizes with platelet count (rs115127268, PP4 = 0.77, PP4/(PP3+PP4) = 0.82). **C)** An eQTL and sQTL for *C12orf4* colocalizes with serum phosphate (rs116827959, PP4 = 0.94). In each panel: QTL is shown on top and GWAS on bottom, lead variant from colocalization analysis is indicated with rsID and set to reference with LD calculated using 1000 Genomes Africans. GWAS are from PAN-UKBB Africans.

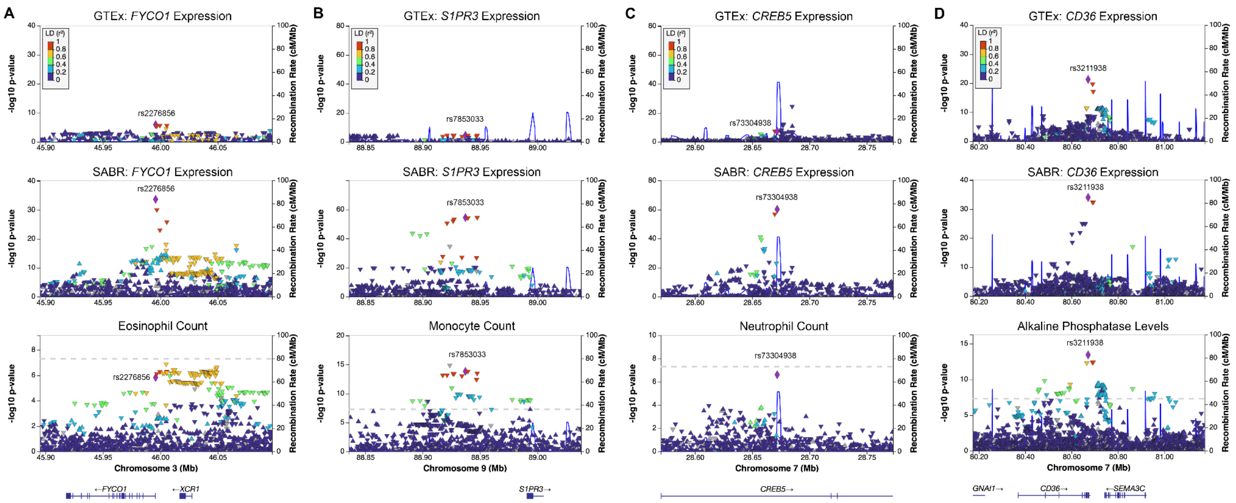

**Figure S17. PAN-UKBB African GWAS colocalizations with African-enriched QTLs having dramatically increased significance in SABR versus GTEx whole blood. A)** An eQTL for *FYCO1* colocalizes with eosinophil count (rs2276856, PP4 = 0.86, GTEx p-value = 1.15e-6, SABR p-value = 2.08e-34). **B)** An eQTL for *S1PR3* colocalizes with monocyte count (rs7853033, PP4 = 1.00, GTEx p-value = 2.91e-5, SABR p-value = 3.8e-55). **C)** An eQTL for *CREB5* colocalizes with monocyte count (rs73304938, PP4 = 1.00, GTEx p-value = 4.16e-8, SABR p-value = 5.54e-61). **D)** An eQTL for *CD36* colocalizes with monocyte count (rs3211938, PP4 = 1.00, GTEx p-value = 5.17e-22, SABR p-value = 9.20e-35). In each panel: GTEx whole blood QTL is shown on top, SABR QTL in the middle, and GWAS on bottom, lead variant from colocalization analysis is indicated with rsID and set to reference with LD calculated using 1000 Genomes Africans. SABR and GTEx y-axis scales have been set equal in each panel to enable comparison. GWAS are from PAN-UKBB Africans.

### Supplementary Methods

#### Study Design

This study involved recontacting individuals from South Africa who had previously contributed their genetic data to the AWI-Gen study at baseline and enrolling them in the SABR study [1–4]. This study was submitted for ethics approval as an independent study and did not include AWI-Gen participants sampled from cohorts in West and East Africa. Phenotype information was previously collected and used during cell type deconvolution analysis [4].

##### SABR Participant Selection

Participant recruitment and sample collection took place in the following Study Centres: The SAMRC/Wits University Rural Public Health and Health Transitions Research Unit which hosts and operates the Agincourt Health and socio-Demographic Surveillance System (HDSS), (Tsonga, also referring to themselves as Shangaan, Xitonga speaking individuals); Developmental Pathways to Health Research Unit (DPHRU, Zulu, isiZulu-speaking individuals), Soweto; and DIMAMO (formerly Dikgale), University of Limpopo (Pedi or Bapedi, sePedi-speaking individuals) [5,6]. Recruitment to constitute participants of the multicentre AWI-Gen cohort involved random sampling of equal numbers of adults from the three established population-based cohorts (Agincourt, DIMAMO, DPHRU) hence reflecting the underlying population composition.  Participants for recontacting were selected based on their self-reported language group membership and genotype data generated from the H3Africa SNP array in the AWI-Gen study [7]. Briefly, individuals were selected based on close clustering in principal component analysis with other members of the same group, and being minimally related (pi-hat < 0.0625). Following community engagement and individual informed consent we enrolled 750 adult participants from three different South Eastern Bantu-speaking groups (approximately 250 from each group and ~ 50% women): Bapedi, XiTsonga, and Zulu. For Tsonga and Bapedi groups, only participants for whom all four grandparents lived at the study location, or had the same self-reported group membership, were eligible for recruitment (in exceptional circumstances, three such grandparents were considered appropriate). Participation was voluntary and potential participants were excluded if they chose not to participate after receiving information about the project. Due to the objectives of the study and limited resources, we could not be inclusive of all community members and therefore had to apply specific inclusion criteria: (1) Individuals who provided informed consent; (2) Individuals whose grandparents (all four or at least three) were identified by the participants as being from the same ethnolinguistic group (Bapedi, XiTsonga, or Zulu); (3) Individuals who were taking any medication that could interfere with the transcriptome analysis or had a condition that could interfere with blood collection; (4) Individuals who were pregnant or lactating, or are otherwise part of a vulnerable population in the context of this study.

##### Sample Collection and Storage

Trained nurses who were supervised and supported by senior staff of each research center collected approximately 6 ml of venous blood in two 3 ml Zymo DNA/RNA shield tubes (catalog

#R1150) from each participant. Study sites collected and stored samples according to standard operating procedures. Centralized sample storage and processing took place at the Sydney Brenner Institute for Molecular Bioscience (SBIMB) at the University of the Witwatersrand in Johannesburg. Laboratory work (DNA and RNA extraction and whole genome sequencing) took place in the commercial laboratory of Psomagen, Inc. (Maryland, USA). The DNA and RNA were destroyed after they had been analyzed in accordance with the informed consent.

#### Community Engagement & Ethics Review

Using established community engagement and information dissemination channels, we engaged community stakeholders through in person meetings (when possible) at each Study Centre before participants were approached and informed consent sought. In the semi-rural rapidly transitioning rural context of Agincourt (Mpumalanga) and the peri-urban environment of DIMAMO (Limpopo) Study Centres, we met with community advisory boards of the HDSS Centres in May 2021 to explain what the study was about, how the selection and consenting process would proceed, and what benefit sharing would entail for their communities. It was an opportunity to hear their views, respond to their questions and concerns, and to adjust our processes in line with their suggestions. We also received feedback on the following issues: (a) How they felt about the project in general and whether they considered that it would be valuable for their communities to participate; (b) If they felt comfortable with how their data would be used and what would happen to the data, for example data sharing with the wider scientific community. It was stressed that no individually identifying data would be shared; (c) How they felt about providing biological samples and disposal and/or repatriation of the samples.

In the urban Soweto study setting, given that this was during the COVID-19 pandemic, our partners interacted with the municipal ward-councilors to explain the study to them and then proceeded with recruitment in line with the processes of the Developmental Pathways for Health Research Unit.

Recruitment started in June 2021 and was completed by November 2021. All three research centers were mindful of taking precautionary measures related to possible infection during the pandemic and used masks and sanitisers during all interactions, with frequent decontamination of transport vehicles (if they were transporting participants) and recruitment study rooms. No adverse events were reported during the recruitment phase.

Once participant recruitment was completed, the community benefit sharing process was put in place and competed by November 2022. This process will be described elsewhere and was in line with the processes developed by Variant Bio [8].

Ethical approval for this study was provided by the Human Research Ethics Committee (Medical) of the University of the Witwatersrand (M210214), University of Limpopo Research Ethics Committee (TREC/199/201:IR) and the protocol was reviewed by the Variant Bio ethics committee. Individual participants gave voluntary informed consent to participate in the study before taking part.

### Sequencing and Data Processing

#### Whole Genome Sequencing, Joint Variant Calling, and Imputation

Whole blood samples were sent to Psomagen (Maryland, USA) for DNA extraction, library preparation and sequencing. DNA samples were prepped with Illumina DNA Prep library kits and sequenced on NovaSeq 6000 instruments with 2x151bp reads targeting throughput of ~16Gb/sample, for an expected deduplicated coverage of about 5x per sample.
WGS data were then processed with Variant Bio's in-house pipeline, which includes extensive quality control and processing following the GATK Best Practices guidelines (CCDG functional equivalence version) [9]. Median coverage among the mid-pass WGS samples was 5.5x, ranging from 3.9x to 17.7x per sample. Following an approach for mid-pass WGS we have previously described, joint genotyping and subsequent imputation was performed across a total of 1,399 genomes by including 100 South African individuals sequenced at high-coverage as part of H3Africa, multiple populations from 1KGP (GBR, YRI, CHB, LWK, GWD, for a total of N=514 individuals), and various Bantu and San populations from the Human Genome Diversity Project and the Simons Genome Diversity Project (N=28 individuals) [10–14]. Before imputation with BEAGLE, all genotypes with GQ < 18 were filtered (set to missing), as well as variant alleles observed only in a single individual removed [15]. In a first pass, we only imputed genotypes of variants in Genome In a Bottle (GIAB) high-confidence regions, ensuring high quality imputation within this stricter set [16]. After fixing these genotypes, we added variants outside high-confidence regions as well as structural variants (HGSVC SVs genotyped in all individuals using PanGenie, with GQ < 200 set to missing) and imputed these as well [17]. Indels in GIAB low-confidence regions were filtered from all downstream analyses.

#### RNA-Sequencing and Expression / Splicing Quantification

RNA extraction and sequencing was done at Psomagen (Maryland, USA). All successfully extracted RNA samples with RIN > 5 and DV200 > 70 were prepped with Illumina TruSeq stranded Total RNA Ribo-Zero Globin kits. The resulting 669 libraries which passed library QC were sequenced on NovaSeq 6000 instruments with 2x151bp reads, targeting 9Gb per sample. Reads were processed with Variant Bio's in-house pipeline following the GTEx/TOPMed RNA-seq pipeline employing RNA-SeQC (v2.3.6) for gene quantification of collapsed genes (GENCODE v34) [18]. Splice junction counts were quantified with the GTEx Leafcutter pipeline (without WASP filtering) [19,20]. Extensive QC was performed including sample identity checks versus the WGS data. A total of 29 samples were removed as a result: 19 with low RNA-seq quality (ribosomal content > 10%, medTIN < 30%, Exonic Rate < 20%, or pct_correct_strand < 90%), 7 where WGS or RNA mismatched previous array data, 3 samples that were sequenced in duplicate.

#### Variant Annotations

Variants genotyped from whole genome sequencing were annotated with the following: Variant Effect Predictor (VEP) using cache version 1.0.8, Combined Annotation Dependent Depletion (CADD) using version 1.6, and allele frequencies calculated from high-pass whole genome sequencing of 1000 Genomes individuals [12,21,22]. In addition, variants were annotated based on their allele frequencies across populations. The following labels were used:

- Globally common: MAF > 5% in all 1000 Genomes populations
- African-enriched: MAF in 1000 Genomes Africans > 5x all other continental ancestries
- African-specific: MAF = 0 in all other 1000 Genomes continental ancestries
- SABR-enriched: MAF in SABR > 5x 1000 Genomes Africans
- SABR-specific: MAF = 0 in 1000 Genomes
- Enriched in each SABR group: MAF > 5x at least one other SABR group
- European-absent: MAF 0 in 1000 Genomes Europeans

##### Genotype Quality Control Filtering for Downstream Analyses

Subject-level and variant-level quality control (QC) metrics derived from genotype data were used to filter inputs for downstream analyses using Hail (v0.2.57). Subject-level exclusion filters included: imputation rate > 85% in the final cohort, heterozygosity > 3 standard deviations from mean, relatedness (pi-hat) > 0.0625, any of PCs 1-5 > 7 standard deviations from mean. Variant-level exclusion filters included: minor allele count < 10 in final cohort, imputation rate > 75% in final cohort, indels in GIAB low confidence regions, variants not in Hardy-Weinberg equilibrium (p < 1e-10), variants not on main autosomes. For QTL analyses, subjects were first subset to only those with RNA-seq data passing QC. For genetic analyses including population allele frequencies of functional variants and xCell GWAS, 724 subjects and 22,569,620 variants passed all QC filters. Of the 724 subjects, 620 had expression data. For QTL analyses, 614 subjects and 17,880,016 variants passed all QC filters.

#### Cell Type Deconvolution

##### Cell Type Deconvolution using xCell

Cell type deconvolution from bulk blood transcriptome data was performed using the xCell method [23]. This is the same method extensively benchmarked and used by the GTEx consortium [24]. The method produces cell type enrichment scores, which are useful for ranking the relative enrichment of a given cell type across samples within the cohort, however the estimate is not directly interpretable or portable across cohorts. This makes it a suitable phenotype for downstream analyses including GWAS and cell type interaction QTL analyses, where the data are inverse normal transformed and only the rank within the cohort is relevant. Cell type enrichments were estimated in a combined cohort of GTEx whole blood alongside SABR samples. This was required in order to benchmark SABR results against GTEx. xCell produces estimates for a variety of cell types, only some of which are relevant to whole blood. To define the cell types used in downstream analyses, we filtered based on those with an enrichment score > 0 in at least 50% of the cohort, and those in the myeloid, lymphoid, or hematopoietic stem cell categories (Table S3). This resulted in the selection of 40 blood relevant cell types.

##### GWAS of Cell Type Enrichment Scores

For blood relevant cell types, inverse normal transformed xCell enrichment scores were used for GWAS with the following covariates: sex, age, age$^2$, sex*age, sex*age$^2$, mean whole genome sequencing depth, and genetic principal components 1-20. A total of 620 participants were included in the GWAS. All variants with minor allele count >= 12 were included in the GWAS. GWAS were run using the linear_regression_rows() function in Hail (v0.2.57). For each GWAS, genomic inflation was calculated using lambda gc ($\lambda_{gc}$) and all GWAS had $0.95 < \lambda_{gc} < 1.05$.

#### QTL Mapping

##### *cis*-QTL Mapping

In order to maximize compatibility with the largest existing QTL resource, the v8 *cis*-QTL mapping pipeline developed by the GTEx Consortium was used [25]. For eQTLs, genes with ≥ 0.1 TPM in ≥ 20% samples and ≥ 6 reads (unnormalized) in ≥ 20% samples were included. Gene expression values calculated using TMM were inverse normal transformed and used as phenotypes for QTL mapping. QTL mapping was performed using fastQTL in a +/- 1MB window from the gene TSS with between 1,000 and 10,000 permutations per gene [26]. Variants with minor allele count >= 10 corresponding to a MAF >= 0.81% were included. Age, sex, genetic principal components 1-20, mean whole genome sequencing depth, and 60 PEER factors inferred from the RNA-seq data were included as covariates [27]. The top permutation derived p-values per gene were corrected for multiple testing using Storey's *q* value and significance was defined at FDR < 5% [28]. For sQTLs, a strategy similar to eQTLs was used. Intron clusters to be included in QTL mapping were defined using the same thresholds as in GTEx v8. The intron usage phenotypes were inverse normal transformed and fastQTL was run in the grouped permutation mode with the "--grp" argument, grouping by the gene each splice event mapped to. PEER factors were inferred from the splice qualifications and 15 were included as covariates, alongside the same covariates as the eQTL mapping. While multiple sQTLs (corresponding to different splicing clusters) were mapped per gene, only the top, most significant sQTL was reported. For downstream analyses, the nominal significance threshold established by permutation mapping could be used to identify sQTLs in addition to the most significant cluster. Multiple testing correction was performed across all genes using the same approach as eQTLs.

##### Independent *cis*-eQTL Mapping

To map independent *cis*-eQTLs a stepwise conditional analysis approach was used. Briefly, for each significant eGene (FDR < 5%) the top eVariant was included as a covariate and the regression was re-run for all variants within the *cis* window. If the variant with the lowest p-value in the step met the gene-level nominal significance threshold determined by permutation mapping it was considered conditionally independent and included along with the preceding variant as a covariate in another round of regression. Subsequent steps were performed until no variant met the permutation mapping nominal significance threshold, up to a maximum of 20 steps. For each gene, the number of conditionally independent signals, and the top variants from each step were reported.

#### *cis*-ieQTL Mapping

A similar ieQTL mapping approach as Kim-Hellmuth *et al.* was used to map interaction eQTLs from xCell enrichment estimates [24]. Briefly, xCell enrichment scores were inverse normal transformed, variants meeting a MAF cutoff in both the top and bottom half of the normalized xCell distribution were tested, and the same PEER factors as for *cis*-eQTL mapping were used as covariates. A nominal pass for interaction mapping was performed with fastQTL using the "--interaction" argument. Gene-level multiple testing correction was performed using eigenMT to estimate the number of independent tests per gene, and then the top variant was selected and its nominal p-value corrected using Bonferroni correction [29]. The Bonferonni corrected top variant p-values were then corrected across all genes using the Benjamani-Hochberg (FDR) approach.

Using neutrophils (the same cell type Kim-Hellmuth *et al*. mapped in whole blood), we benchmarked how changing three parameters impacted the number of ieQTLs mapped: minor allele frequency cutoff (0.025 or 0.05), residualizing xCell estimates on age and sex before inverse normal transformation (yes or no), and the variance explained threshold in eigenMT (0.99, 0.975, 0.95). The results of the benchmarking are shown in the table below.

| MAF Cutoff | Sex / Age Regressed | Variance Explained | Median Tests* | ieGenes FDR < 5% | Comments |
|---|---|---|---|---|---|
| 0.05 | No | 0.99 | 3,080 | 55 | Kim-Hellmuth approach |
| 0.05 | Yes | 0.99 | 3,078 | 58 | |
| 0.025 | No | 0.99 | 4,251 | 63 | |
| 0.025 | Yes | 0.99 | 4,249 | 69 | Most ieGenes at 99% var |
| 0.05 | No | 0.975 | – | 72 | |
| 0.05 | Yes | 0.975 | – | 71 | |
| 0.025 | No | 0.975 | – | 87 | Most ieGenes at 97.5% var |
| 0.025 | Yes | 0.975 | – | 86 | |
| 0.05 | No | 0.95 | 902 | 89 | |
| 0.05 | Yes | 0.95 | 902 | 89 | |
| 0.025 | No | 0.95 | 1,347 | 97 | |
| 0.025 | Yes | 0.95 | 1,348 | 104 | Most ieGenes at 95% var |

* median tests per gene from eigenMT. Note that this was not recorded for the 0.975 variance runs.

Based on the results, we decided to use the approach that maximizes the number of ieGenes mapped - a MAF cutoff of 0.025 and a more liberal variance explained threshold of 0.95 for eigenMT. Benchmarking of eigenMT by Davis *et al.* found that using a variance threshold of 0.95 is slightly less conservative than permutation mapping, whereas 0.99 is more conservative than permutation mapping. This caveat should be noted for the ieQTLs mapped in this study.

Regressing out age and sex from xCell scores had an inconsistent and marginal effect on neutrophil ieGene discovery. However, no sample had an xCell neutrophil enrichment score of "0", where residualization could help break ties before inverse normal transformation. To further investigate, we benchmarked the effect of regressing age and sex out of xCell enrichment scores before residualization for a cell type with a high number of "0" estimates. To this end, we ran CD4+ T cell ieQTL mapping (N samples with 0 score = 139) with a MAF cutoff of 0.025 and eigenMT variance threshold of 0.95 with and without residualization. We found that residualizing before transformation reduced the number of ieGenes mapped (shown in table below). Therefore, we decided not to include this in our ieQTL mapping approach.

| **MAF Cutoff** | **Sex / Age Regressed** | **Variance Explained** | **Median Tests** | **ieGenes** FDR < 5% | **Comments** |
|---|---|---|---|---|---|
| 0.025 | No | 0.95 | 1,350 | 16 | More ieGenes |
| 0.025 | Yes | 0.95 | 1,350 | 12 | |

Due to a lack of full summary stats available from Kim-Hellmuth *et al*. we were not able to test SABR neutrophil ieQTLs for replication in GTEx whole blood. However, we were able to test GTEx ieQTLs for replication in SABR. A notable caveat is the very different LD structure between the cohorts, with likely overall shorter haplotypes in SABR versus GTEx due to African ancestry. This would be expected to lower replication of GTEx ieQTLs in SABR, but not vice-versa. Despite this, we observed significant correlation in the interaction term betas (Spearman rho = 0.18, p = 4.40e-6) and moderate evidence of replication (Storey's pi1 = 0.30) for 622 significant GTEx neutrophil ieQTLs (FDR < 1%) with ieVariant MAF > 5% in SABR. Notably, replication improved dramatically when using ieQTLs with a more stringent FDR (Figure S15).

#### Colocalization Analysis

PAN-UKBB GWAS summary statistics in GRCh38 coordinates from African individuals (XX_AFR) were downloaded from the Broad Institute [30]. The full path to the summary statistics for each of the GWAS used for colocalization is listed under path_sumstats in Table S15. Colocalization analysis was performed using the *coloc* R package (version 5) under a single causal variant assumption with variant minor allele frequencies and p-values as inputs [31]. Loci were selected for colocalization based on having a GWAS variant with p < 5e-6 within +/- 500KB from the lead QTL variant, for all significant QTLs (FDR < 5%). Two thresholds for colocalization were used to capture the maximum number of putative colocalizations; lenient: PP4 > 0.50 and PP4 / (PP3 + PP4) > 0.80; strict: PP4 > 0.80. When visualizing colocalization loci, the reference was set to the lead colocalization variant and LD was calculated using 1000 Genomes Africans.
